# Superspreading Events Without Superspreaders: Using High Attack Rate Events to Estimate N_º_ for Airborne Transmission of COVID-19

**DOI:** 10.1101/2020.10.21.20216895

**Authors:** Mara Prentiss, Arthur Chu, Karl K. Berggren

**Affiliations:** Department of Physics, Harvard University; Department of Electrical and Engineering and Computer Science, Massachusetts Institute of Technology

## Abstract

We study transmission of COVID-19 using five well-documented case studies – a Washington state church choir, a Korean call center, a Korean exercise class, and two different Chinese bus trips. In all cases the likely index patients were pre-symptomatic or mildly symptomatic, which is when infective patients are most likely to interact with large groups of people. An estimate of *N*_0_, the characteristic number of COVID-19 virions needed to induce infection in each case, is found using a simple physical model of airborne transmission. We find that the *N*_0_ values are similar for five COVID-19 superspreading cases (∼300-2,000 viral copies) and of the same order as influenza A. Consistent with the recent results of Goyal *et al*, these results suggest that viral loads relevant to infection from presymptomatic or mildly symptomatic individuals may fall into a narrow range, and that exceptionally high viral loads are not required to induce a superspreading event [1,2]. Rather, the accumulation of infective aerosols exhaled by a typical pre-symptomatic or mildly symptomatic patient in a confined, crowded space (amplified by poor ventilation, particularly activity like exercise or singing, or lack of masks) for exposure times as short as one hour are sufficient. We calculate that talking and breathing release ∼460*N*_0_ and ∼10*N*_0_ (quanta)/hour, respectively, providing a basis to estimate the risks of everyday activities. Finally, we provide a calculation which motivates the observation that fomites appear to account for a small percentage of total COVID-19 infection events.

## Background and Motivation

Growing evidence supports the idea that aerosols - particles of diameter less than 5-10 µm that can remain suspended in air for many hours - are an important channel of spread of COVID-19 [3,4]. The SARS CoV-2 virus has a diameter of only 60-140 nm [5], so an aerosol of diameter 1 µm has the same volume as of order 1,000 virions. Further, aerosols can stay suspended for long periods: in still air, it takes a 1 µm diameter particle about 12 hours, and a 3 µm particle about 1.5 hours to vertically settle 5 feet [6]. When considered by number (not by total volume), droplets emitted when coughing, breathing, or speaking are nearly all of order 1 µm [7–10]. An index patient who emits infective aerosols should therefore leave particles with the capacity to fit hundreds or thousands of virions suspended in the air for many hours. Recently, viable SARS-COV-2 virus was recovered from aerosols in the rooms of COVID-19 patients, providing the first direct evidence of potential aerosol induced infection [11], and the US CDC has also now officially recognized aerosols as a potential transmission mechanism [12].

Interestingly, most COVID patients do not infect anyone, whereas a small number of patients infect multiple people [2,13,14]. The observed distribution of attack rates for COVID-19 cannot be obtained if all encounters are assumed to have the same virus transmission probability; however, if some encounters have a higher virus transmission probability than others, then some index patients may pass the virus to several people, even though most index patients do not pass the virus to anyone. We consider two factors that might increase the transmission probability for some encounters: 1. an index patient who emits unusually large quantities of live virus (superspreader) 2. a venue that accumulates live airborne virus emitted by an average index patient (enhanced transmission). If one person infects many people during a single event, that interaction is often described as a superspreading event, which could be explained either by the presence of a superspreader or by enhanced transmission.

Support for superspreaders has been provided by measurements of viral loads across all stages of disease progression which indicate that viral loads span ∼8-9 orders of magnitude centered on ∼ 10^6^ -10^7^ copies/mL [15–19]. Support for the role of superspreaders was also offered by studies of the Skagit choir that inferred an index patient viral load ranging from 10^9^ - 10^11^ copies/mL [20,21], which is at the very high end of measured viral loads.

Support for “enhanced transmission” is provided by measurements and modeling from Goyal *et al*, which indicate that the period of peak infectivity for COVID-19 is ∼0.5-1.0 days, coinciding with a relatively narrow range of peak viral loads (measured by nasopharyngeal swab) of order 10^7^ copies/mL [1,2]. Goyal *et al* considered the known distribution for the rate of secondary infections due to patients with COVID-19. That distribution includes both patients who do not transmit the virus to anyone and those who transmit it to many people. They then fit that distribution to a model of transmission to obtain the parameters for the model that best characterize COVID-19 transmission. The results of the fits indicate that superspreading events occur when an index patient has a large number of contacts during the period of peak viral load. In addition, they find that, compared to influenza patients with an equivalent physical contact network, patients with COVID-19 infect a greater number of contacts (which they attribute to aerosolization.) Monte-Carlo modeling also suggests that superspreader events result from “enhanced transmission” due to airborne transmission of virus emitted by presymptomatic patients [22].

To address the question of whether superspreading events require superspreaders from a different point of view, we consider the following well-documented events that have appeared to involve aerosol transmission of SARS-CoV-2:

1. In January 2020, an index patient who rode a 100-minute round trip bus ride to a Buddhist worship event in China in January, infecting 23 out of 67 bus riders. Conveniently, there was a second “control group” bus which went to the same event, in which 0 out of 60 riders were infected [23].
2. Also in January 2020, an index patient in Wuhan, China took two different buses on a single trip. On the first 2.5 hour leg on a larger bus, 7 of 48 riders were infected, including riders ∼4.5 m away; on the second 1-hour leg on a smaller bus, 2 of 12 riders were infected [24].
3. In February 2020, clusters of coronavirus transmission in aerobic dance exercise classes were found, in Korea, this time due to sick instructors [25]. These classes were very short – only about 50 minutes – and 5-22 students were in a ∼60 m^2^ ∼ 650 square foot room (so that many were presumably separated from the instructor by more than 2 m.
4. In March 2020, 94 out of 216 employees working on the same floor of a large (∼1,000 m^2^) Korean call center tested positive. Of the positive results, 89 of them were one one side of the floor measuring over 400 m^2^ [26].
5. Also in March 2020, 61 singers convened for 2.5 hours of choir practice in a ∼200 m^2^ church in Skagit County, Washington, in the U.S., after which 32 were confirmed positive and 20 were suspected positive [20,27].

Past work has estimated the number of infective quanta emitted per unit time and their implications for infection risk. For example, Miller *et al* and Buannono *et al* have analyzed the Skagit Choir case in detail, [20,22] while Jimenez has published an online tool using the findings to evaluate aerosol infection probabilities in different settings [28]. More recently, Bazant has published estimates of source strengths due to various activities and introduced a framework for interpreting and calculating the subsequent risks [21].

## Overview

In this paper, we use the above five case studies to directly estimate the characteristic number of virions *N*_0_ needed to induce infection, assuming a probability of infection after inhalation of *N* virions which varies as 1 – exp(-*N/N*_0_). In each of these cases, there is no measurement of viral loads of the index patients; however, if for each case we assume the viral density characteristic of peak infectivity (10^7^ copies/mL, as measured by nasopharyngeal swab [1,2]), we find a value of *N*_0_ ranging between 322-2,012 copies, which is similar to values assumed by other authors for COVID-19^1^ and the ∼300-9,000 copy infectious aerosol dose for Influenza A [32]. Thus, our derived *N*_0_ values in combination with previous experimental measurements [1,2] support the following: (1) none of these five so-called “superspreading” events required super spreaders; (2) the distribution of viral loads in index patients at the times most relevant for airborne transmission is narrow compared to the total distribution of viral loads in the population; (3) if that narrow value is coincident with measured loads at the onset of symptoms, then *N*_0_ values are of the order of levels found for influenza and (4) that airborne transmission can be enhanced by accumulation of viral particles in a confined space, which allows an index patient with an average viral load to infect many other people.^2^

We review the existing literature to gather estimates of as many of the key parameters involved in aerosol transmission of COVID-19 and use these estimates to infer the *N*_0_ for SARS-CoV2. Figure 1 summarizes the main components of this calculation, starting with the viral load in the index patient and concluding with the estimated probability of infection. For each of the major variables, we show the overall ranges in the literature, as well as highlighting (in red) the input values used for the point estimate of *N*_0_. We discuss all of the major parameters in the text below, but highlight two points here: first, there is a ∼3-4 order of magnitude difference in the literature between the volume of assumed aerosol emission across difference measurements, which has a direct bearing on the question of whether superspreading events require index patients with exceptional viral loads; second, that despite the very wide range of viral loads (∼9 orders of magnitude), the final values of *N*_0_ across the cases are remarkably consistent and within ∼1 order of magnitude. While the absolute values of *N*_0_ will change depending on the aerosolized volume assumption, this consistency would hold for any single value of the viral density chosen.

**Figure 1:**
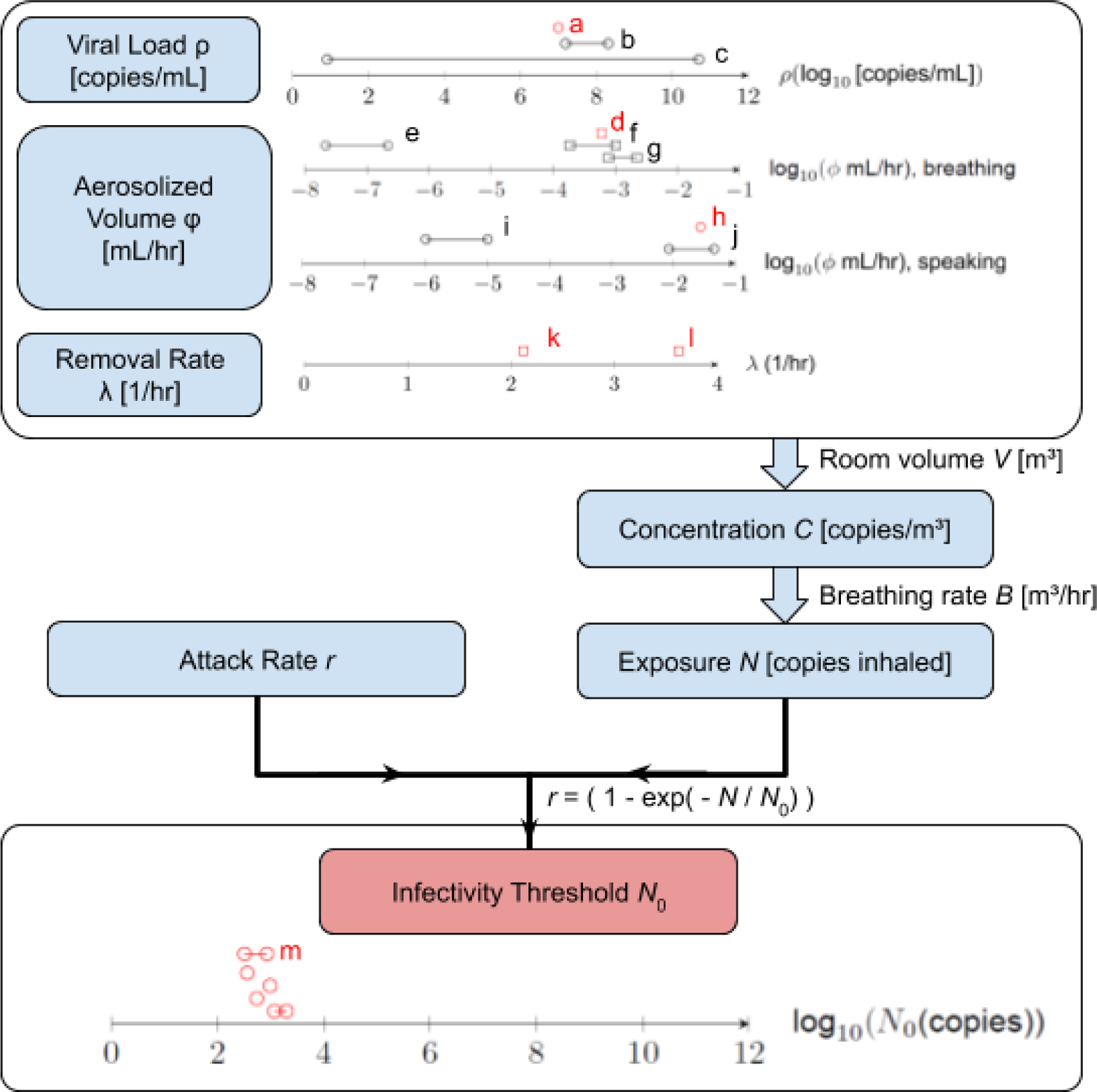
Overview of Key Parameters. For each parameter, we show the value(s) used in this work in red and key ranges: the viral density *ρ*, the aerosolized volume emitted per unit time ϕ, the removal rate λ, the exposure time *T*, and the room volume *V* determine the concentration C of virions in the room, which determines the number of virions inhaled given the breathing rate *B*. The attack rate *r* (infected / exposed) is used to infer *N*_*0*_. Density *ρ* (Log_10_ Copies/mL): (a) value used (10^7^ copies/mL); (b) range observed by Kleiboeker *et al* across 4,432 patients[15]; (c) modeled peak viral load found by Goyal *et al* across 25 patients[1]. Aerosolized volume ϕ (mL/hr): <u>Breathing</u>: (d) value used (6.0 × 10^−4^ mL/hr); (e) range in Miller *et al* based on Morawska *et al* [10,20]; (f) range in Stadnytskyi *et al* based on Morawska *et al* [10,33]; (b) range based on exhaled breath condensate (EBC) dilution measurements [34–36]; <u>Talking</u>: (h) value used (2.7 × 10^−2^ mL/hr); (i) range in Miller *et al* based on Morawska *et al* [10,20]; (k) range in Stadnytskyi *et al* [33]; Removal rate λ (1/hr) (k), (l) value used for cases in buildings and buses, respectively (2.12 h^-1^ and 3.62 h^-1^) (m) Resultant point estimates for *N*_*0*_. Each line or circle represents a range or value estimated from a case: from top to bottom, U.S. (Skagit) choir; Korean call center; Korean fitness center; China Buddhist bus trip; Wuhan bus trips. The reasoning behind the choice of value used in this work is described in detail in the main text.

Independent of the estimate of *N*_0_, and as Miller, Jimenez, Buannano, and Bazant have separately done, the case studies can be used to compute the number of aerosolized units of *N*_0_ (quanta) released per unit time, which can in turn be used to estimate the quanta released by talking or breathing [20–22,28]. This calculation finds that talking releases ∼460 *N*_0_ per hour of aerosolized virus, whereas normal breathing releases ∼10*N*_0_ per hour. We use these estimates to provide some order of magnitude risk estimates for everyday activities. Because we associate these quanta emission rates with typical peak viral loads, we find that these quanta are a reasonable representation of population-wide risk of aerosols in various scenarios. It should be noted, however, that if, as has been suggested in other analysis [20,21], the case studies such as the Skagit choir correspond to individuals with a viral load of 10^9^ - 10^11^ copies/mL (as opposed to the ∼10^7^ copies/mL in this analysis), the population-wide risk will be much lower than that implied by the quanta calculations (since such high viral loads are in the 95+ percentile of measured loads). Finally, we present a calculation indicating that typical fomite exposures might be on the order 1-10 virions, which may motivate the observation that only a small percentage of total transmission appears to be through fomites [37].

## Exposure Model

A common dose response model (Wells-Riley model) assumes that it takes order *N*_0_ virions in to infect a person [38]. Specifically, each virion is assumed to be independent of the others and to have a probability 1/*N*_0_ of infecting an individual. If the individual is exposed to *N* particles, the expected number of infective events is *N*/*N*_0_. Treating the infection as a Poisson process, the probability of zero infective events (i.e. the probability of not being infected) is exp(-*N*/*N*_0_) and the probability *p*(*N*) of being infected is 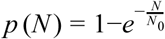.

In this section, we estimate both *N* and *N*_0_ for the five cases mentioned above – the Skagit Choir in the U.S., the Korean call center, the Korean aerobics class, and the two Chinese bus trips. In particular, we quantify the key source (viral density and expelled aerosol volumes), removal (air exchange and inactivation/settling), and exposure parameters (breathing rate, time, room volume) based on analysis of the current literature.

If we know the concentration (virions/unit volume of air) of virions *C*(*t*) and that a person inhales the air at a constant breathing rate *B* for a total time *T*, the total number *N* of virions breathed in is

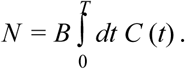

The value of the breathing rate *B* has been well-characterized in a number of studies, and is typically assumed to be in the range of 0.45 m^3^/hr - 0.60 m^3^/hr for sedentary activities [30,39]. Relevant for this analysis is the measurement of Binazzi *et al*, which found that quiet breathing, reading with a normal voice, and singing all had similar breathing rates (0.54 ± 0.21 m^3^/hr, 0.54 ± 0.21 m^3^/hr, and 0.61 ± 0.40 m^3^/hr, respectively) [40]. The breathing rate rises to ∼1.3-1.5 m^3^/hr during “moderate” activities(easy cycling, climbing stairs) and ∼2.5-3.3 m^3^/hr during “heavy” activities (cross country skiing, climbing with load)[41]. For purposes of this analysis, we use *B* = 0.5 m^3^/hr for the choir, call center, and bus cases, and *B* = 2.0 m^3^/hr for the fitness center case.

The concentration *C* depends on both spatial coordinates and time. It is common in indoor air quality calculations to make the simplifying assumption that the air is “well-mixed” – i.e. that the virions are spread evenly across the volume in the room so that one can neglect the spatial dependence. This assumption is justified if the time for virions to spread across the room is small compared to the timescale of the decay processes above due to air exchange and other virion losses. The mixing time for cough-generated aerosols has been characterized, where a coughing simulator was placed on one end of a 2.7 m × 2.7 m × 2.4 m environmental chamber, and the concentration of small aerosols (0.3-4.0 µm) was measured as a function of time at various locations [42]. The measurement found that after about 5 minutes, the concentration reached a steady state which was uniform across the room. Note also that in the experiment, the chamber was sealed during the coughing simulation (zero air exchanges per hour) so that there was no airflow that would cause additional mixing. These results support the reasonableness of making the well-mixed approximation.

With the well-mixed approximation, we can calculate the number of virions *n*(*t*) over time. We assume that the person emits a constant flow *S* of new virions per unit time, and that since the air is well-mixed, the number of virions lost to decay per unit time is λ·*n*(*t*). There are multiple possible sources of decay so λ is the sum of multiple contributions λ_1_ = λ_1_+ λ_2_ + λ_3_… which will be specified below, so *n*(*t*) obeys

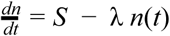

which has the usual solution of a transient term decaying at a rate λ toward a steady state *n*_eq_:

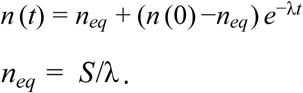

In the cases we consider, we assume that the environment starts off “clean” – i.e. that n(0) = 0. That is, we assume there are no viral particles in the air before the choir assembles, or before the riders enter the bus, etc. In this case, we have

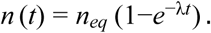

Since the concentration *C* = *n* / *V* where *V* is the volume of the room, we can solve for the total inhaled particles *N* over a time *T* (where B is the breathing rate i.e. the volume inhaled per unit time).

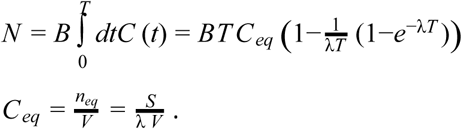

To perform the calculations for the five case studies, we need to know *B*, the breathing rate; the room volume *V* (either reported or estimable); the exposure time *T* (generally reported); the decay rate λ; and the source term *S*. We now turn to the last two terms.

### Decay rate λ

As noted above, λ is the sum of several contributions, namely:

λ_1_= air exchange rate = rate at which fresh air replaces stale air in the room. For example, if λ_1_ = 2.0 h^-1^, the air turns over twice an hour in the room. See the Supplementary Materials for representative values of λ_1_.

λ_2_ = decay rate of SARS-CoV-2 in air, due to both settling and inactivation in air. For settling, Diapouli *et al*. report aerosol settling rates in indoor settings which average approximately 0.3 h^-1^: PM 2.5 decay rates ranged from ∼0.1-0.4 h^-1^, while measured PM 10 decay rates range from ∼0.05-0.65 h^-1^ [43]. For SARS-CoV-2 viral deactivation, both van Doremalen *et al* and Fears *et al* measured the decay in infective virus in a Goldberg drum, which in principle eliminates the effects of settling. However, van Doremalen measured a half-life of SARS-Cov-2 1.09 hours in air (λ_2_ = 0.64 h^-1^) [44], while Fears *et al* were not able to detect meaningful viral inactivation of 2-3 μm SARS-CoV-2 aerosols over a 16 hour period (i.e. measured λ_2_ = 0) [45].^3^ For purposes of this calculation, we include a contribution of 0.3 h^-1^ due to settling, and a contribution of 0.32 h^-1^ due to inactivation (averaging the van Doremalen and Fears measurements), for a total value of λ_2_ = 0.62 h^-1^.

λ_3_ = effect of filtration. In the case studies considered, there is no filtration, so we will return to this in the scenario analysis.

### Source term *S*

We estimate the source term as the product of a viral density *ρ* (virions / unit volume) and a volume per unit time of aerosol expelled *ϕ*.

### Viral density *ρ*

This quantity has been directly measured, but it ranges over many orders of magnitude across patients. Across a sample of 3,303 patients testing positive in Germany, Jones *et al* measured a mean viral load of 5.72 Log_10_ copies/mL with an estimated standard deviation of ∼1.9 Log_10_ copies/mL [18]. Similarly, in a sample of all 4,428 positive RT-PCR from a single US laboratory tests (defined as a cycle threshold value for RT-PCR ≤ 38.0), Kleiboeker *et al* measured a mean and median, respectively, of 5.85 and 6.05 log_10_ copies/mL, with a range of 0.91-10.42 Log_10_ copies/mL, 15.3% results greater than 8 log_10_ copies/mL, and an estimated standard deviation of ∼2.0 log_10_ copies/mL (Figure 1) [15]. ^4^ Other measurements include Arnaout *et al* (initial positive results from 4,774 patients, mean of ∼5.2 log_10_ copies/mL) and Jacot *et al* (initial positive results from 4,172 patients, mean of ∼6.5 log_10_ copies/mL, and median of 6.77 log_10_ copies/mL) [16,17]. Arnaout finds a maximum viral load of 2.5 × 10^9^ copies/mL (9.4 log_10_ copies/mL) while Jacot finds a maximum viral load of ∼2 × 10^10^ copies/mL (10.3 log_10_ copies/mL), broadly consistent with Kleibocker and Jones. In all of these measurements, the probability of a given viral density drops sharply beginning at ∼10^8^ copies/mL.

We note two additional points about these distributions. First, the measurements show only total viral copies derived from cycle threshold (Ct) values, without making any statement about whether these copies were infectious. Second, the remarkable breadth of the distribution -- spanning 8-9 orders of magnitude in viral titre -- needs to be interpreted carefully. The PCR tests underlying the distribution were taken at different points after infection, so the wide range of viral loads arises from not only inhomogeneity across patients, but also, changes during disease progression.^5^ Said differently, a single patient tested continuously from exposure to recovery would show a range of viral loads from the limit of detection (∼100 copies/mL) to the peak viral load (conceivably 10^7^-10^9^ copies/mL).

As we discuss in detail later, there is generally a narrow time period of maximum infectiousness, estimated by Goyal et al to be approximately 0.5-1.0 day around the time of peak viral load [2]. Based on the viral trajectory of 16 hospitalized patients, Goyal et al estimated a peak viral load, as measured by nasopharyngeal (NP) swab, of ∼7.2-7.3 Log_10_ copies/mL. For purposes of this analysis, we therefore use a value of *ρ* =10^7^ copies/mL, which approximates the average viral density in the window of ∼1 day around the peak. This is also the value that was used in Monte Carlo modeling of the individual infection risk for susceptible subjects confined in a room with an asymptomatic index patient [22]. As a side note, this value is also not far from the mean density measured by Wölfel *et al*, who found a mean viral density in sputum of 7.00 × 10^6^ viral copies per mL (6.85 Log_10_ copies/mL) [19]. The assumed value of *ρ* is the ∼66th percentile of values measured by Kleiboeker *et al*, as shown in Figure 2.

**Figure 2:**
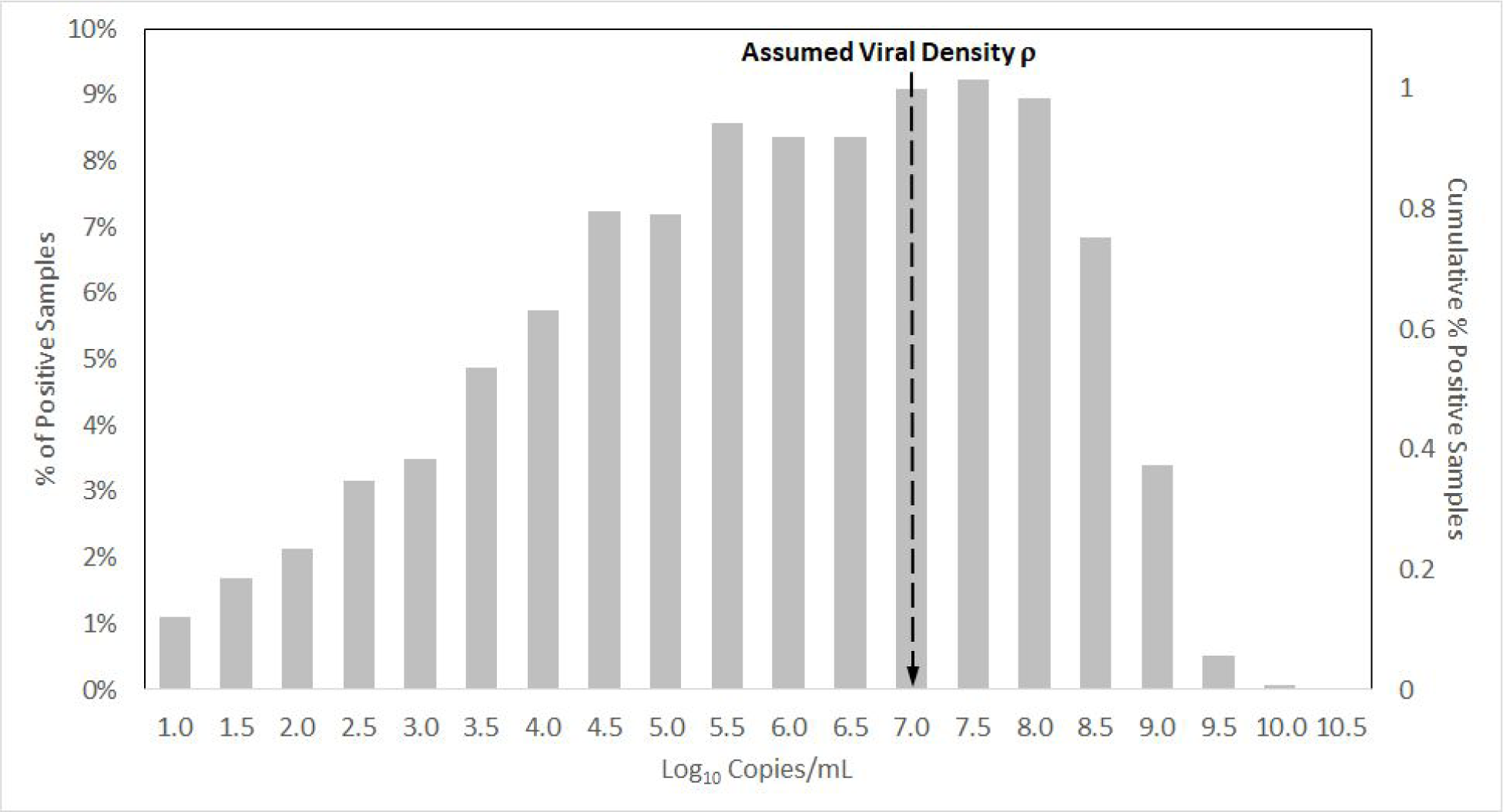
Viral Load Distribution (Kleibocker *et al*.) and Assumed Viral Density *ρ*. Viral load distribution (copies/mL) as measured by Kleibocker *et al*. Viral loads were derived from all 4,428 positive test results (cycle threshold value for RT-PCR ≤ 38.0) in a U.S. testing laboratory; values include all viral copies, some of which may not be infective. Viral load is referenced to the left hand axis and shows a peak near 7.5 log_10_ copies/mL, near the assumed value of *ρ*. The cumulative sum of the Kleibocker et al. distribution is also shown, in reference to the right hand axis.

### Volume expelled per unit time *ϕ*

The value of *ϕ* (volume of aerosols expelled per unit time) depends on what the person is doing. We use measured values for speaking and then infer the results for other activities such as talking, breathing, and singing.

#### Talking

Stadnytskyi *et al* recently measured aerosol volumes by scattering of laser light off droplets generated during repetition of the phrase “stay healthy” for 25 seconds [33]. The study infers a dehydrated droplet diameter of ∼4 μm and a hydrated droplet diameter ∼12-21 μm, corresponding to hydrated volumes of 60 to 320 nL. We use the average of these two values – i.e. when talking, *ϕ* = 190 nL/25s = 7.6 nL/s.

#### Breathing

Morawska *et al* measured differences in expelled particle density when breathing and talking (e.g. saying “aah” and counting) [10]. Averaging across the talking and breathing scenarios, these measurements show that that talking releases about an order of magnitude more particles by number than does breathing (ratio of ∼18.1×), but also, that because vocalization releases larger particles, the volume ratio of talking to breathing is higher, at ∼45.7×.^6^ Thus, for breathing we use *ϕ* = 1/45.7x 7.6 nL/s ∼ 0.2 nL/s.

#### Singing

Singing expels about 6 times as many particles as talking [9,47], so for this case we use *ϕ* = 6 × 7.6 nL/s = 45.6 nL/s.

The order of magnitude of the volume of aerosols emitted during speaking varies significantly in the literature depending on the measurement method used. Specifically, Miller *et al* estimate, based on the work of Morawska, that speaking results in emission of ∼1-10 nL of aerosols per hour [10,20]. In contrast, Stadnytskyi *et al* find volumes of 60-320 nL over just 25 seconds of speaking -- some 3-4 orders of magnitude higher. This large difference may be due partially to the differences in particle sizes measured. Stadnytskyi *et al* estimate that the aerodynamic particle sizer (APS) method used by Morawska *et al*. may measure particles of hydrated diameter ∼8.7 μm and less, which are outside of the 12-21 μm range in the scattering experiment [33]. Nonetheless, a sharp drop of 3-4 orders of magnitude for a halving of the particle diameter would not be expected. A second factor that may account for the difference is that the measurement of Morawska *et al* uses an APS, which may not count all particles emitted. Asadi *et al*., who perform similar measurements to Morawska using an APS, note that their reported particle emission rates are to be viewed in relative, not absolute, terms [9].

We believe that Stadnytskyi’s estimate is the more appropriate one to use here. As a cross-check to this estimate, we can compare our computed value of *ϕ* when breathing to experiments that measured respiratory fluids using exhaled breath condensates (EBCs). In an EBC measurement, a patient breathes into a chilled tube which collects the condensed breath. That condensed breath can then be tested for volatile and non-volatile solutes, including pathogens [35,36,48].^7^ Breath samples consist almost exclusively of water vapor from the lungs. The water vapor cannot carry the virus, but the breath samples also contain a small contribution of aerosols from respiratory fluids. Those aerosolized fluids can carry viruses, as well as other non-volatile condensates that are present in the respiratory system. Thus, EBC’s include both condensed water vapor and condensed aerosols. Effros *et al*. compared the concentration of non-volatile solutes in condensed respiratory fluids to the concentration of the same solutes in airways. They found that the concentration of the non-volatile solute in EBC’s was reduced by an average factor of ∼20,000:1 (± ∼2,500:1) over the concentrations in lungs, with the reduction factor varying between ∼1000:1 to ∼50,000:1 [34].^8^ If the non-volatile solute concentration in exhaled droplets is similar to concentrations in lungs, then respiratory aerosols account for ∼1/20,000 of the total water loss through breathing. Water loss through breathing is well-characterized, at ∼350-400 mL/day (∼16 mL/hour) [36,50]. The dilution factor of 20,000 from EBC measurements would therefore imply aerosolized breath volumes of 16/20,000 = 8 × 10^−4^ mL/hour, consistent with our value of *ϕ*_breathing_ (∼6 × 10^−4^ mL/hour.)^9^ In a separate publication, Effros *et al* estimate that ∼4.5 nL of airway lining fluid is exhaled per liter of breath [35], which, assuming a breathing rate of 500 L/hour (0.5 m^3^/hour), translates into ∼2.3 × 10^−3^ mL/hr of emitted volume, within a factor of ∼4 of our figure for *ϕ*_breathing_ of ∼6.0 × 10^−4^ mL/h. That is, the volume of breathing aerosols implied by Stadnytskyi’s speaking measurements and our assumed ratio of breathing/speaking volumes are consistent with the volume of breathing aerosols from EBC measurements.

To further motivate why particles of a few μm diameter, may constitute the main contribution to aerosol spread of infection, we consider the measurements of Asadi *et al*, who showed that speech droplets follow an approximately lognormal distribution peaking at diameter ∼1 μm, the fit of which is shown as the dashed line in Figure 3 below. In general, the infectivity of airborne pathogens will depend on their deposition locations in the body, which in turn depends on particle size: for instance, particles of ≲ 3 μm can reach lung tissue, whereas larger (≳ 8 μm) particles may lodge in the upper airways [51]. Furthermore, viral densities (or the density of viable virions) may depend on particle size. (It should be noted that Santarpia *et al*. successfully recovered infectious SARS-Cov-2 from particles of less than 4.1 μm, including particles with diameters < 1μm [52]. They note, however, that failure to recover infectious SARS-Cov-2 from particles larger than 4.1 μm may have been an artifact of the effect of the collection mechanism on viral viability.) We assume for simplicity that over the relevant size range, both the viral density and the aerosol infectivity are independent of the aerosol volume, in which case what matters for infectivity is not the number of particles of each size but rather their volume contribution. For purposes of estimating the relative volume contributions of particles of different sizes to transmission, we multiply the particle number distribution by two factors: (i) the volume contribution of each diameter *d* (proportional to *d*^3^) and (ii) a factor accounting for the longer lifetimes of smaller particles in air. For simplicity, we assume that each particle size has a characteristic survival time τ given by the Newton-Stokes law (τ = 4.5 η *h* / *g ρ r*^2^ where η is the viscosity of air, *h* is the height, and *r* = *d*/2 is the radius, *ρ* = density of water), and that the number of particles after a time *t* is proportional to exp(-*t*/τ).[30,53] The solid line in Figure 2 shows that the relative volume contribution for *h* = 2m and *t* = 1 hour (the timescale of removal of particles at typical air exchange rates) peaks around 3 μm.

**Figure 3:**
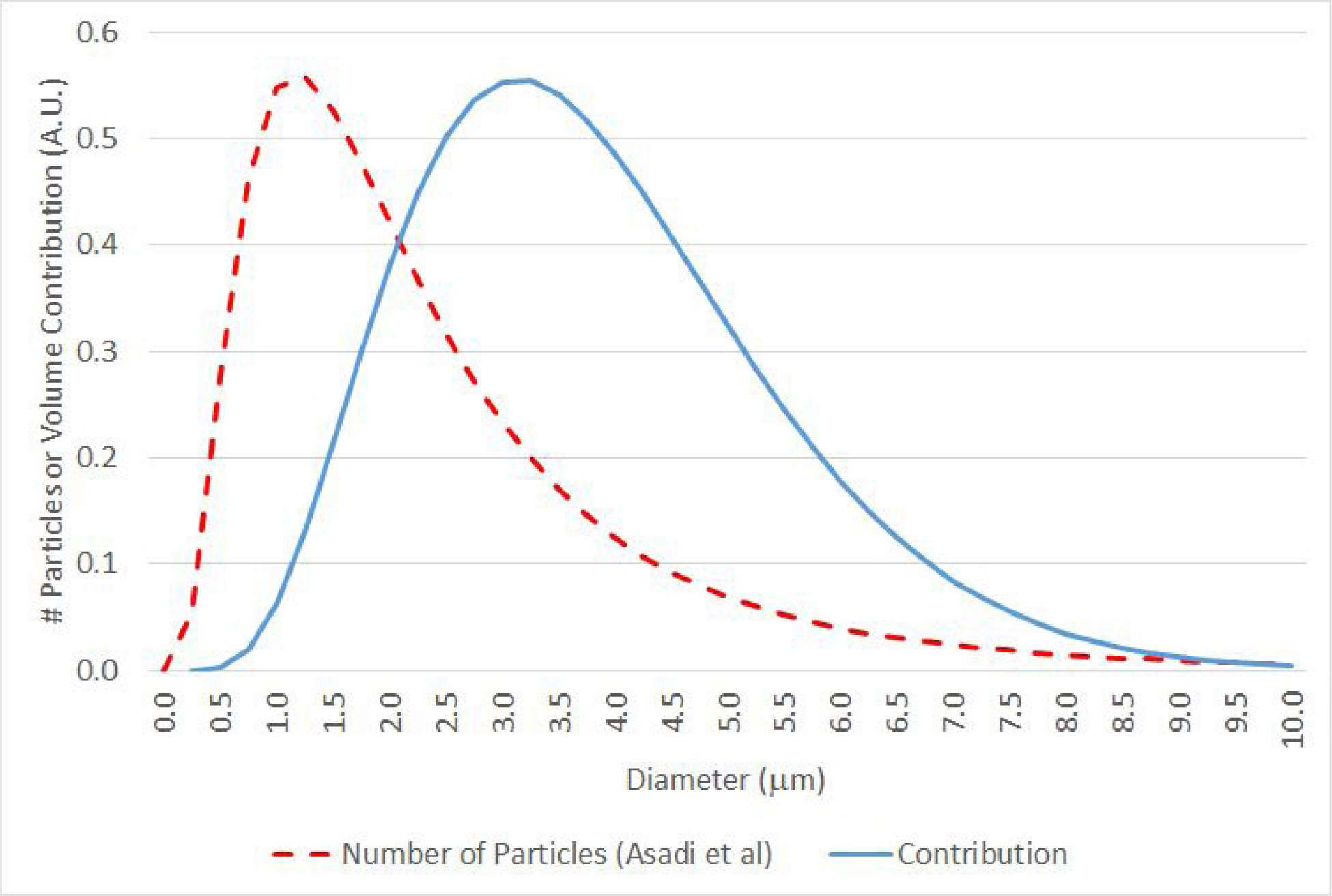
Number of Aerosol Particles and Relative Volume Contribution by Diameter. Number of aerosol particles (dashed line) and relative contribution adjusting for particle volume and airborne lifetime. The number of aerosol particles at each diameter is reproduced from Asadi *et al*. (Figure S3), which uses an aerodynamic particle sizer to measure the fully dehydrated particle size distribution after a 5 second vocalization [9]. The relative contribution adjusting for particle size multiplies the particle number distribution by a factor proportional to the volume of each particle (∼diameter^3^) and a factor proportional to particle airborne lifetime (∼exp(-const·diameter^2^)). These observations suggest that particles in the range of 4 μm diameter may be the primary contributors to infectivity.

### Typical Value of *ρ*

Putting these together, we get *S* = *ρ ϕ* for various activities: Talking: *S* ∼ 4,600 virions/minute; breathing: *S* ∼ 100 virions/minute; singing: *S* ∼ 27,000 virions/minute.

## Estimating Characteristic Dose (*N*_0_) Using Case Studies

In each of the five case studies, an attack rate or range of attack rates (number of infected/ number of exposed) *r* is reported along with a description of the environment. From this information, along with the source and decay parameters discussed in the previous section, *N* and therefore *N*_*0*_, can be estimated from *r* = 1 - exp (*-N/N*_*0*_).

In estimating *N*, the physical parameters of the case studies are not all known with precision and it is necessary to make assumptions about many of them, particularly the air exchange rate. Here, we use the EPA office building assumption of 1.5 h^-1^ for the choir, office, and fitness center cases, and 3.0 h^-1^ in the bus case.^10^ ^11^ In addition, given the large difference in virion expulsion for different activities, we must make an assumption about the percentage of time spent talking/singing versus breathing. For choir singing, based on published observations, we assume that ⅔ of the time is spent singing [54]; in the cases of the call center, fitness class, and buses, respectively, we assume that 90%, 50%, and 25% of the time are spent talking.^12^ Finally, there can be ambiguity in the attack rate: in the case of the Skagit Choir, there were 32 confirmed infected and 20 probable infected, so the attack rate ranges from 53-87%; we calculate *N*_0_ in both cases [27]. In the case of the Korean Fitness center, the reported attack rate across all events is 26% (57 confirmed/217 exposed); however, this includes an instructor teaching yoga and pilates (0 confirmed/25 exposed), which we exclude from the analysis [25]. Finally, in the case of the Korean call center, out of 94 total cases on the 11th floor, 89 of them were in an area separated from the rest of the floor by elevator banks and other rooms [26]; we calculate the attack rate for this area, but show that the value of *N*_0_ is similar if one instead uses the entire area.

Table 1 shows the key inputs and outputs for the calculations. For the interested reader, we include in the Supplementary Materials a spreadsheet where these inputs can be customized. For the base inputs, the range of *N*_0_ (322-2,012) that results from the five scenarios is reasonably narrow. The order of magnitude of *N*_0_ calculated here is not unreasonable. It has been speculated that *N*_0_ is of order a few hundred to a few thousand particles for SARS-CoV-2 [31].

**Table 1:** Key Inputs and Outputs for the Case Studies. (Refer to the spreadsheet in the Supplementary Materials for details). The quanta/hour (scenario average) refers to the quanta emission rate averaging the high and low attack rate cases, for the particular parameters of that scenario. The quanta/hour (talking, breathing) are the implied quanta/hour for talking and breathing.

|  | <u>Skagit</u><br><u>Choir</u> | <u>Korean call</u><br><u>center</u> | <u>Korean</u><br><u>fitness</u><br><u>center</u> | <u>Buddhist</u><br><u>Bus</u> | <u>Wuhan</u><br><u>(Bus 1)</u> | <u>Wuhan</u><br><u>(Bus 2)</u> |
| --- | --- | --- | --- | --- | --- | --- |
| **Inputs** |  |  |  |  |  |  |
| Relevant activity | Singing | Talking | Talking | Talking | Talking | Talking |
| % of time in activity | 67% | 90% | 50% | 25% | 25% | 25% |
| % of time breathing | 33% | 10% | 50% | 75% | 75% | 75% |
| Source term S (virions/h) | $1.1 \times 10^6$ | $2.5 \times 10^5$ | $1.5 \times 10^5$ | $7.2 \times 10^4$ | $7.2 \times 10^4$ | $7.2 \times 10^4$ |
| $\lambda_1$ (air exchange rate $\text{h}^{-1}$) | 1.5 | 1.5 | 1.5 | 3.0 | 3.0 | 3.0 |
| $\lambda_2$ (settling + inactivation $\text{h}^{-1}$) | 0.6 | 0.6 | 0.6 | 0.6 | 0.6 | 0.6 |
| Total decay rate $\lambda$ ($\text{h}^{-1}$) | 2.1 | 2.1 | 2.1 | 3.6 | 3.6 | 3.6 |
| Total time T (h) | 2.5 | 8.0 | 0.8 | 1.7 | 2.5 | 1.0 |
| Breathing rate B ($\text{m}^3 / \text{h}$) | 0.5 | 0.5 | 2.0 | 0.5 | 0.5 | 0.5 |
| Volume of Space V ($\text{m}^3$) | 810 | 1,143 | 180 | 50 | 71 | 34 |
| Attack rate (infected /population) | 53%-87% | 65% | 30% | 34% | 15% | 17% |
| **Outputs** |  |  |  |  |  |  |
| Viral copies/$\text{m}^3$ (time averaged) | 519 | 96 | 207 | 276 | 254 | 428 |
| Viral particles inhaled during T | 649 | 384 | 345 | 230 | 317 | 214 |
| **$N_0$** | **322-851** | **361** | **980** | **547** | **2,012** | **1,175** |
| Quanta / hour (scenario average) | 2,347 | 683 | 152 | 133 | 36 | 62 |
| Quanta / hour (talking) | 586 | 757 | 279 | 500 | 136 | 233 |
| Quanta / hour (breathing) | 13 | 17 | 6 | 11 | 3 | 5 |

As a further reasonableness check, we calculate airborne viral copy concentrations in a range of 96-519 copies/m^3^ across the scenarios. These can be compared with measurements of Liu *et al*., who measured viral densities at various locations in a Wuhan hospital (a location with filtration and higher air exchange) and found densities of ∼10-40 copies/m^3^ in some staff areas (range 0-42), and up to 113 copies/m^3^ intensive care units (range 0-113) [55]. Hallway air samples in the University of Nebraska National Quarantine Unit (NQU, where mildly ill patients are isolated), found detectable virus in 58.3% of cases, ranging from ∼2,000-9,000 copies/m^3^ [56].

We also show the quanta/hour emitted in each scenario (the number of *N*_0_ emitted by the index patient in each of the case studies), and the implied quanta/hour emitted by speaking (range 136-757, average of 461) and breathing (range 3-17, average of 10). These quanta are broadly consistent with those found by other authors: Miller *et al* find an emission rate of 970 quanta/h for the Skagit choir case [20];^13^ Bazant finds ∼4-16 quanta/h for breathing and 54-970 quanta/h for speaking and singing [21]. Jimenez, based on the work of Buonnano *et al*, recommends a range of ∼2-14 quanta/hour for oral breathing (depending on the activity) and 61-408 quanta/h for loud speaking [22,28].

In the attached spreadsheet, we also show the case in which the entire Korean call center is considered (greater area and lower attack rate), which results in a similar *N*_0_ = 240.^14^ In the Supplementary Materials, we also discuss the case of a restaurant in Guangzhou, which is included in the spreadsheet but not in the case studies due to measurement ambiguities in both the attack rate and the relevant room volume. The values of *N*_0_ for the restaurant case are broadly consistent, however, with the five cases here, with a range of *N*_0_ of 499 to 2,415.

Though SARS-CoV-2 and influenza A infections have many dissimilar features, it is interesting to note that the values of *N*_*0*_ (∼300-2,000 copies) here are similar to that of influenza. Our search of the literature finds that most measurements would indicate an *N*_*0*_ range of ∼300-9,000 viral copies for influenza (by way of comparison, Killingley and Bischoff quote a range influenza *N*_*0*_ of ∼350-1,700 and ∼130-2,800, respectively [57,58].) In the case of influenza A aerosol infection, the human ID_50_ (dose at which 50% of subjects become infected) is widely cited as 0.6-3.0 TCID_50_ (the viral load at which 50% of cells in culture are infected) [59]. However, the range of viral copies per TCID_50_ varies significantly and has been measured by Fabian *et al* [60], Ward *et al* [61], and Yang *et al [32]* to be 300, 1000, and 452-2,100 viral copies, respectively. That is, in the case of influenza A, the human ID50 is in the range of ∼200-6,000 copies, which would imply a *N*_0_ (=ID50/*ln*(2)) for influenza of ∼300-9,000 copies [62].

As a further comparison, Fears *et al*., measured the change in infectivity of aerosolized SARS-Cov-2 with time. That work reports both genome copies per unit volume and PFU per unit volume [45]. Over 5 measurements spanning ∼ 8 hours, the ratio of PFU to genome copies ranges from ∼20 to ∼1,100, with an average of ∼500 copies/PFU.^15^ If, as in the case of influenza, the ID50 for inhalation of SARS-CoV-2 is on the order of 1 PFU, this measurement would imply an ID50 of ∼500 copies, so that *N*_0_ = 500/*ln*(2) ∼ 700, in accord with our range of *N*_0_ of 300-2,000.^16^

The consistency of the *N*_*0*_ estimates from the 5 cases -- each of which was derived assuming the same viral density -- is surprising in light of the extremely broad range of viral loads discussed previously (spanning ∼8-9 orders of magnitude). If, in the alternative, we assume that *N*_0_ is in the range of 100-1,000 and calculate *N*, and thus the viral load, for each scenario, the range of viral loads that result is 5.7-7.5 Log_10_ copies/mL, again much narrower than the total range of viral loads seen in the population (∼2-10 Log_10_ copies/mL). This suggests that the index patients in the five case studies carried viral loads in the ∼ 50-80% percentiles -- above average, but not exceptionally so. More importantly, this consistency suggests that for the types of cases studied here -- where an asymptomatic or mildly symptomatic person infects numerous others through airborne transmission -- the range of viral loads may be much narrower than the range implied by population-wide measurements.

The relatively narrow range of relevant viral loads for “superspreading” events (whether through aerosols or other forms) is consistent with previous epidemiological analyses. Specifically, it is well-established that patients with COVID-19 are most infectious within a window (<∼ 1 day) of the onset of symptoms [37,64]. Goyal *et al* explored this phenomenon quantitatively in a set of two recent papers. [1,2]. In the first, the authors use longitudinal viral load data from 25 patients (i.e. repeated measurements on the same patient) to fit a compartmentalized viral load model. An example set of data and model fit (patient S14) from their work are shown in Figure 3(a) below: the model shows a very rapid increase of ∼5 orders of magnitude over a few days to a peak viral load of order 10^7^ copies/sample, a sharp drop of ∼2 orders of magnitude over the next two days, and a somewhat slower decline thereafter. However, in the window of ± 1 day around the peak viral load, the modeled load varies by only a factor of 10 (∼6.3-7.3 Log_10_ copies/sample) [1].

In the second analysis, Goyal *et al* carry over the viral load model to analyze transmission dynamics in the population. This model finds that the time period in which infected patients are most likely to infect others (more precisely, to have a greater than 50% chance of infecting others), lies within a narrow 0.5-1.0 day period centered around the time of peak viral load [2].

Thus, there should be a significant difference between the viral loads that matter for transmission, which sample only a short time period around the peak viral load, and the viral loads measured by Kleiboeker and others, which sample all potential times after exposure. While the measured viral loads in the 25 longitudinal patient studies varies from ∼100 viral copies/sample (the approximate limit of detection [15,17]) to ∼10^9^ copies/sample, the 25 models find only a very narrow range of fitted peak viral loads of ∼7.2-8.3 Log10 copies/sample.^17^ Goyal *et al* note, however, that, because there is more data during period of viral decline, their fits are generally more reliable in this period [1,2]. Alternatively, Figure 4(b) shows the distribution of the 29 instances of measured (not modeled) viral loads which fall within ± 1 day of Goyal *et al’*s modeled peak viral load (black bars), superimposed on the population-wide viral load measurement of Kleiboeker *et al* (grey bars) [2,15]. While this distribution of actual data is not as narrow as the range of modeled peak distributions, it is clearly far narrower than the population-wide distribution; ∼70% of the data lies in the range of 6.0-7.5 Log_10_ copies/sample.^18^ Interestingly, median viral loads for COVID 19 are consistent with those of a number of other respiratory viruses. Jacot *et al*, for example, reports a median value of 6.77 Log_10_ copies/mL for SARS-CoV2 across 4,326 samples and compares them to 6,050 RT-PCR tests taken over 2015-2020 for 14 other respiratory viruses. All but two viruses show median viral loads between 5.76 and 6.83 Log_10_ copies/mL; for instance, influenza A and B have respective median loads of 6.01 and 6.83 Log_10_ copies/mL [16].^19^

**Figure 4a:**
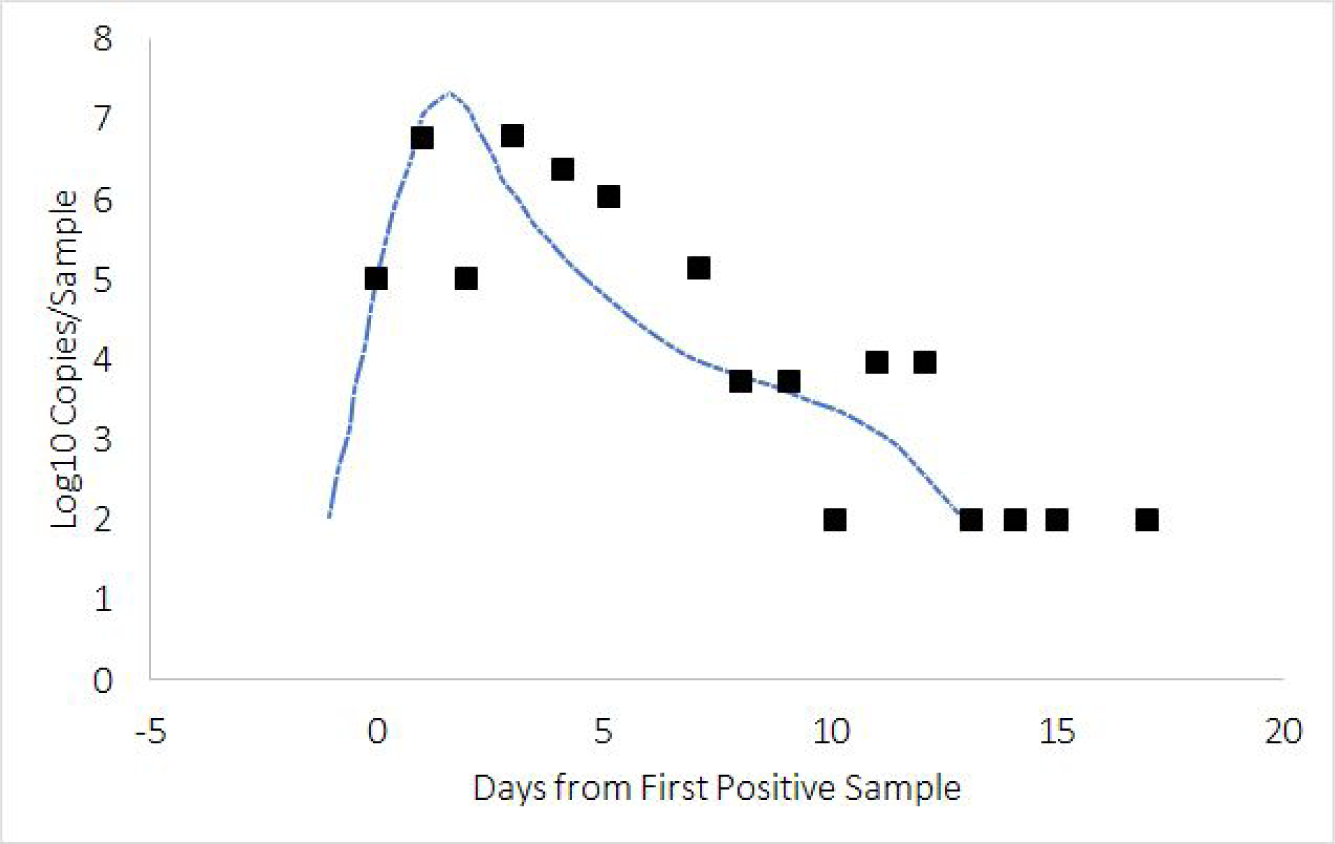
Example Measured and Fit Viral Load Over Time (data and model shown is taken from Patient S14 from Goyal *et al*).[1] The actual and modeled viral loads vary between ∼2 and 7 Log10 copies/sample across the entire period, but the modeled viral load for this patient lies in a narrower range of ∼6.3-7.3 Log10 copies/sample in the period which is ± 1 day around the time of peak viral load.

**Figure 4b:**
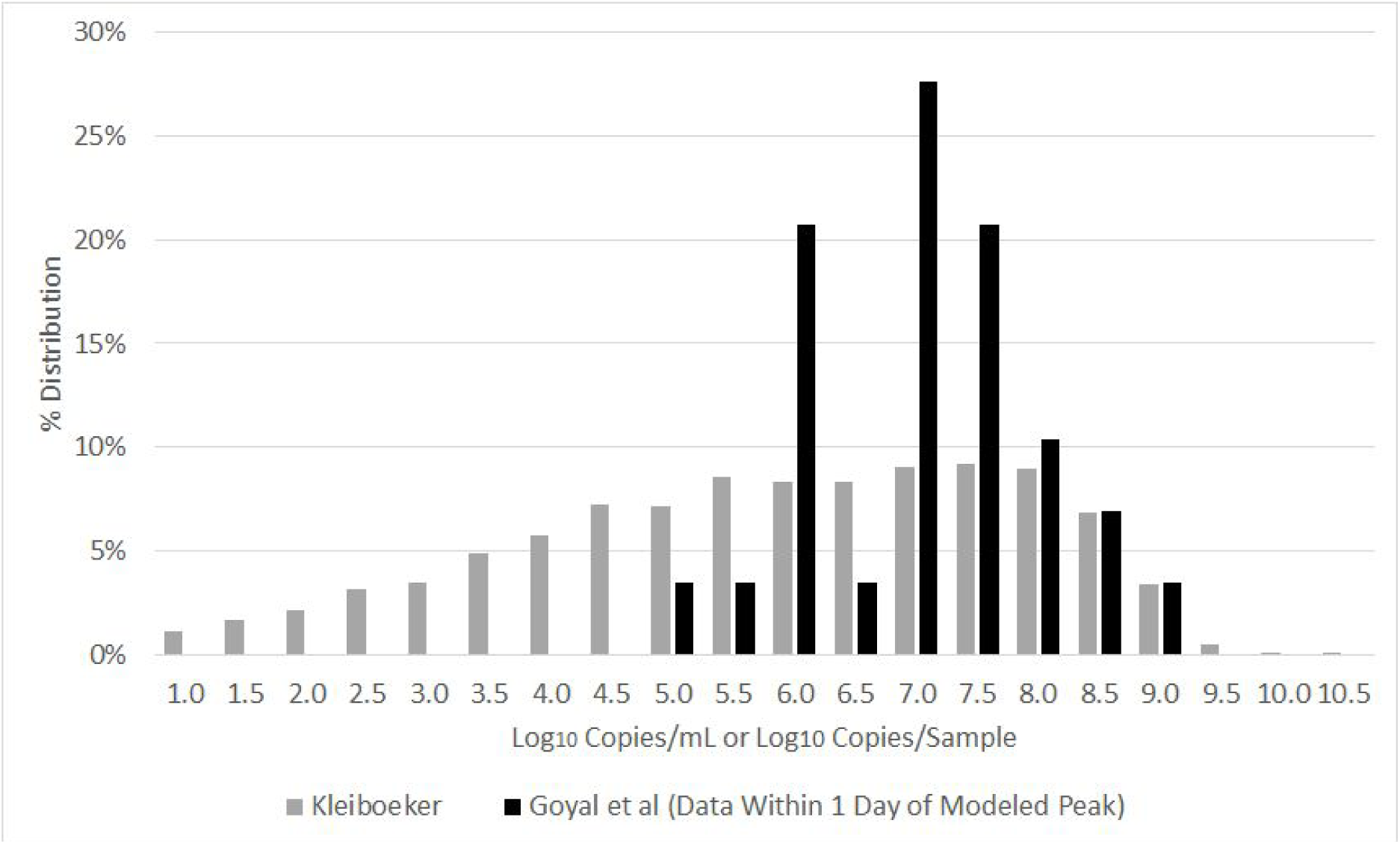
The grey bars are the distribution of viral loads (copies/mL) measured by Kleiboeker et al. The black bars are the distribution of viral loads (copies/sample) drawn from the 25 calibration datasets of Goyal et al, but only for the 29 data points taken within ± 1 day of the modeled peak viral load. The distribution of peak values, which may be more representative of viral load in index patients of our case studies, is significantly narrower than the Kleiboeker distribution.

In sum, the period most relevant for infectivity samples only a narrow slice of the total range of viral loads observed across disease progression, helping to explain why selecting a single viral load value for the five cases results in a modest range for *N*_0_ values (∼300-2,000).

## Risks of Everyday Activities

In a narrow sense, the value of *N*_0_ is not necessary for calculation of the risks of a given situation: one needs only the quanta emitted per unit time. However, in calculating the aggregate risk across the population, it is important to consider whether the calibration quanta emission rates calculated here originated from index individuals with an exceptionally low viral load, or a more typical viral load or an exceptionally high viral load. (By “calibration quanta emission rates”, we mean the quanta per hour emitted by the index patients in the case studies.) Miller *et al* calculate that, assuming that 1 quanta (1 *N*_0_) is equal to 1000 viral RNA copies, the ∼1000 quanta/hour estimated in the Skagit choir case would correspond to a viral load of ∼10^11^ copies/mL [20]. Buonanno similarly calculates that a quanta emission rate of 142 quanta/h, assuming *N*_0_ = 50, corresponds to a viral load of 10^9^ copies/mL (so that if *N*_0_ = 1000, the viral load would be on the order of 10^10^-10^11^ copies/mL [29].) However, in this work we associate similar quanta emission rates based on a viral load on the order of only 10^7^ copies/mL, which implies higher population-wide risks of aerosol transmission.

For example, consider a stylized case of a high-risk activity, where, based on the calibration quanta rates calculated from the case studies (e.g. 460 quanta/hour for speaking, 10 quanta/hour for breathing), the risk of infection given the environmental conditions is 1/3 (i.e. exposure of *N*=0.41 *N*_*0*_). The details of the environment are not important for this purpose; rather, the environment and the quanta together are assumed to give rise to an infection probability of 1/3. Suppose that these calibration quanta emission rates correspond, as found here, to an index patient with a viral density of 10^7^ copies/mL. Across the population, the entire range of viral densities of ∼10^1^ - 10^10^ copies/mL will be represented, each with a different probability of causing infection. An individual with a viral density of 10^6^ copies/mL would, under the same environmental conditions, produce an exposure of 0.041 N_0_ and a risk of infection of 1-exp(−0.041) ∼ 4%; similarly, an index patient with a viral density of 10^8^ copies/mL would produce an exposure of 4.1 N_0_ and a risk of infection of ∼98%.

The population-wide risk of this particular activity assuming a random index patient can be estimated by weighting these differing infection probabilities by the distribution of viral loads present in infected individuals. If the calibration quanta rate corresponds to a viral density 10^7^ copies/mL, and the risk of infection for a person with this viral density is 33%, then the population-wide risk (averaging across the distribution of Kleiboeker *et al*) is calculated to be 30.8%. That is, if the viral density for the calibration quanta is ∼10^7^ copies/mL, the quanta based calculation would be a good representation of the population-wide risk of this activity. On the other hand, if the calibration quanta rates were derived from an index patient with a viral density of 10^9^ copies/mL, the population wide risks (again averaging across the distribution of Kleiboeker *et al*) would be only 2.9% -- an order of magnitude lower -- because relatively few individuals have viral densities as high as 10^9^ copies/mL.

To underscore this point, Table 2 shows the population-wide risks for the activity for different values of the viral density corresponding to the calibration quanta rate, assuming an infection risk of 1/3 for the calibration quanta rate. The table shows the percentile of the viral density distribution for each value of the viral density using the distribution of Kleiboeker *et al* (e.g. 7.0 Log_10_ copies/mL = 71st percentile), the population-wide risk of infection (e.g. 30.8%), and the ratio of this population-based risk of infection to the risk calculated based on the quanta (e.g. 30.8%/33% ∼ 0.92). Previously, we showed that a range of N_0_ of 100-1,000 would, in the five case studies, imply a viral load of 5.7-7.5 Log_10_ copies/mL. Across this range, the order of magnitude of population wide risks (21.9%- ∼50%) is similar to the quanta based risk of 33%. However, if the calibration quanta rates arise from high viral load individuals (10^9^-10^10^ copies/ mL), as has been suggested in earlier works, the population-wide average risk is only 0.4%-2.9%.

**Table 2:** Population Wide-Risks of Infection Assuming Quanta-Based Risk of 1/3, For Different Values of the Viral Density Corresponding to the Calibration Quanta Rate. The risk of infection is assumed to be ⅓ at a given viral density (copies/mL) and then averaged across the distribution of viral densities measured by Kleiboeker *et al* to derive the population-wide risk of infection.

|  | <u>$\text{Log}_{10}$ copies/mL @ Calibration Quanta Rate</u> |  |  |  |  |  |  |  |  |  |
| --- | --- | --- | --- | --- | --- | --- | --- | --- | --- | --- |
|  | <u>5.5</u> | <u>6.0</u> | <u>6.5</u> | <u>7.0</u> | <u>7.5</u> | <u>8.0</u> | <u>8.5</u> | <u>9.0</u> | <u>9.5</u> | <u>10.0</u> |
| Percentile of viral density distribution (%) | 45.2 | 53.6 | 61.9 | 71.0 | 80.2 | 89.1 | 96.0 | 99.4 | 99.9 | ∼100 |
| Population-wide risk of infection (%) | 56.4 | 48.2 | 39.6 | 30.8 | 21.8 | 13.5 | 6.9 | 2.9 | 1.1 | 0.4 |
| Ratio to quanta-based rate of infection | 1.69 | 1.45 | 1.19 | 0.92 | 0.65 | 0.40 | 0.21 | 0.09 | 0.03 | 0.01 |

In analyzing the risks of everyday situations, in addition to the usual input parameters of breathing rate, volume, and so on, we make a few global assumptions:

- *Source strength*: we use the averages from the case studies of 10 quanta/h for breathing and 460 quanta/h for talking.
- *Mask effect*: For mask efficacy, we assume that masks block 71% of outgoing viral particles [65] and 50% of incoming particles, so that masks worn by both infected and susceptible individuals reduce the overall inhaled viral particles by ∼85%.
- *Filtration effect*: assume that we have a filter that exchanges the air at a rate *R* (i.e. passes the air in the room through the filter at *R* times per hour), and that each pass through the filter reduces the amount of the virus by a factor *Q* < 1. For example, if a filter removes 90% of virions on each pass, *Q* = 0.1; a HEPA filter, which removes 99.97% of particles, has *Q* = 0.0003. In the limit that the filter airflow is weak and the room remains well-mixed at all times, the filter is equivalent to adding an amount *R* (1 - *Q*) to the air exchange rate and λ_3_ = *R* (1 - *Q*). However, if the filter airflow is sufficiently powerful that the filtration time is small compared to the mixing time, then after time *t*, the number of times through the filter is *R t* and the viral reduction factor is *Q*^*R t*^ = exp(*R* ln(*Q*) *t*) = exp(-λ_3_ *t*) -- i.e. λ_3_ = -*R* ln(*Q*) [30]. ^20^

The details of each scenario are shown in the spreadsheet in the Supplemental Materials, and the summary risks are shown in Table 3. In cases with small numbers of people (e.g. office, classroom), we assume a single index patient; in cases with large numbers of people (grocery, airplane), we assume a number of index patients dependent on an infection rate and the venue capacity. In each case,we present the probability of infection assuming the index patient(s) are only breathing, only talking, and a mix of breathing and talking (column “scenario”) -- for instance, in an office, talking 15% of the time. Note also that for the cases of the good/bad office/classroom, we assume the same air exchange rate for the “good” and “bad” cases (1.5 or 1.9 h^-1^); the difference in risk is due only to the use of masks and filtration. The absolute risks in which one infected is assumed present are, of course, overstated, since the probability of an asymptomatic patient present would be low, e.g. as of the time of this publication, the U.S. infection rate is ∼1% [66]. However, assuming the presence of an index patient, scenarios with even a small proportion of unmasked/unfiltered speaking time (e.g. dinner party, bad office/classroom) present substantial risks; large volumes (grocery store) or small volumes with strong filtration and masking (commercial flight) are low risk; a case such as a taxi ride, with a short interaction time but a very small (∼3m^3^) volume, is of intermediate risk. We also emphasize that these calculations take into account aerosol transmission only and attribute no risk to direct contact through large droplets or fomites, which may be significant in cases of high density such as a commercial flight.

**Table 3:**
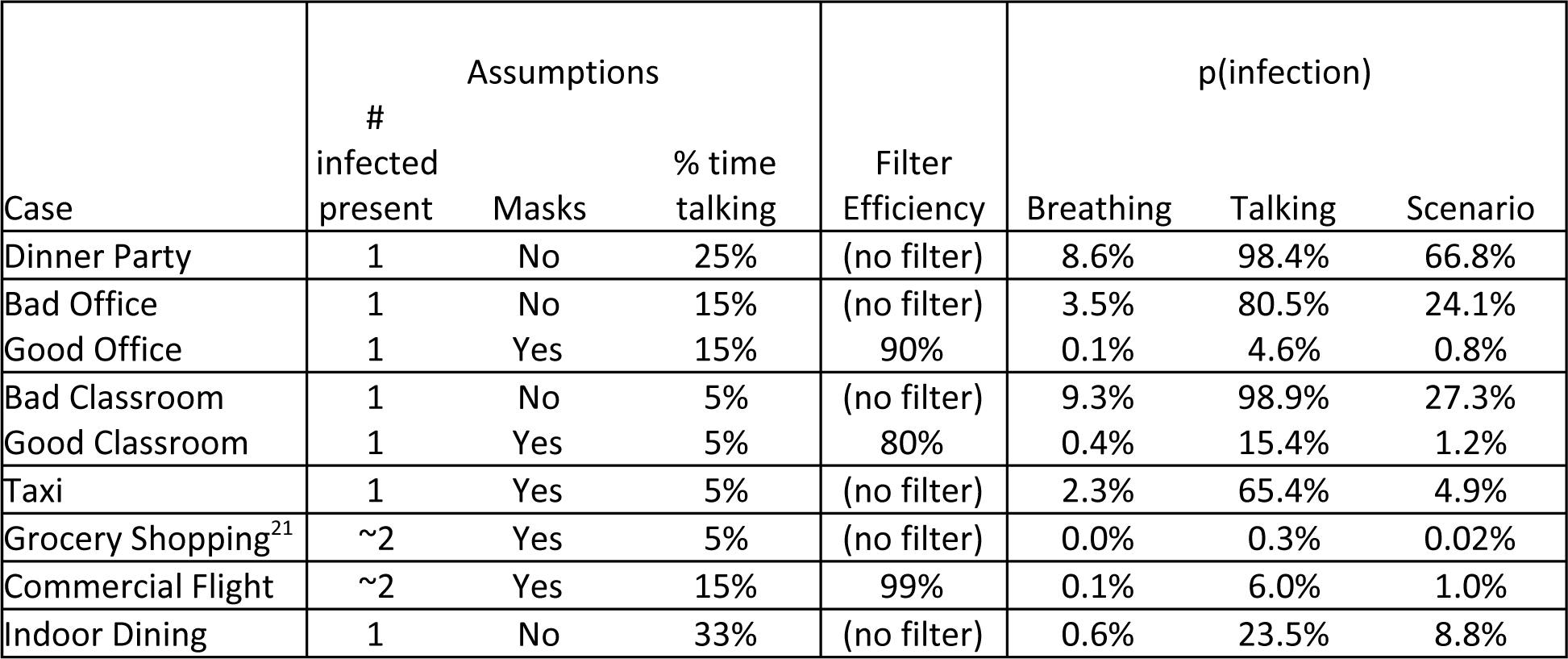
Summary Risks by Scenario. Each scenario assumes a certain percentage of time breathing versus talking, as well as the use, or not, of masks and filters. See the Supplemental Materials for the detailed assumptions of each scenario; these assumptions can be changed by the interested reader.

| Case | Assumptions |  |  | Filter Efficiency | p(infection) |  |  |
| --- | --- | --- | --- | --- | --- | --- | --- |
|  | # infected present | Masks | % time talking |  | Breathing | Talking | Scenario |
| Dinner Party | 1 | No | 25% | (no filter) | 8.6% | 98.4% | 66.8% |
| Bad Office | 1 | No | 15% | (no filter) | 3.5% | 80.5% | 24.1% |
| Good Office | 1 | Yes | 15% | 90% | 0.1% | 4.6% | 0.8% |
| Bad Classroom | 1 | No | 5% | (no filter) | 9.3% | 98.9% | 27.3% |
| Good Classroom | 1 | Yes | 5% | 80% | 0.4% | 15.4% | 1.2% |
| Taxi | 1 | Yes | 5% | (no filter) | 2.3% | 65.4% | 4.9% |
| Grocery Shopping<sup>21</sup> | $\sim 2$ | Yes | 5% | (no filter) | 0.0% | 0.3% | 0.02% |
| Commercial Flight | $\sim 2$ | Yes | 15% | 99% | 0.1% | 6.0% | 1.0% |
| Indoor Dining | 1 | No | 33% | (no filter) | 0.6% | 23.5% | 8.8% |

### Special Case: Fomites

Although substantial effort is expended on disinfecting surfaces, the risk of fomite transmission for SARS-CoV-2 does not appear to be that large. One study estimates that contamination (presumably due to surfaces) accounts for only 6% of total transmission of COVID-19 [37]. The number of virions required to induce infection through fomites (e.g. by touching a contaminated surface and then touching some mucosa such as the nose, eyes, or mouth) may be quite different than the *N*_0_ estimated here for the aerosol pathway. It is nonetheless interesting to estimate the number of virions that might be transferred on occasional touching of public surfaces such as handrails or delivered packages. Zhang *et al* performed a detailed Monte Carlo simulation of an office environment to quantify pathways of influenza transmission [67]. As in Zhang, we assume a sneeze generates 4.75 × 10^−2^ mL of fluid, which corresponds to ∼475,000 viral copies at a viral density of 10^7^ copies/mL. These copies are assumed to be spread uniformly over a circle of radius ∼1 foot (∼3000 cm^2^). The hand (∼25 cm^2^) touches approximately 1% of this area, but, as Zhang notes, only about 7% of virus on a non-porous surface will transfer to the hand (the figure for a porous surface is 3%). Further, about 1 in 4 hand-face contacts is with a mucous membrane, and 35% of the contacts with the membrane successfully transfer virus [67]. Combining all of these factors, the original virus copies are attenuated by a factor of ∼5 × 10^−5^, so that ∼25 virions are ultimately transferred to mucous membranes assuming transfer immediately after the sneeze. Further, van Doremalen et al. measured SARS-Cov-2 half lives of 3.5 and 6.8 hours on cardboard and plastic, respectively, that would further attenuate the transferred virus [44]. In the case of a cough, Zhang estimates a volume emitted of 6.15 × 10^−3^ mL, which would imply transfer of only a few virions [67]. Assuming no great decrease in the scale of *N*_0_, this dose of order 1-10 virions would be unlikely to induce infection.^22^

## Conclusions and Suggestions for Future Research

We estimated *N*_0_, which determines the probability of infection for a person exposed to *N* virions, by applying a simple model of aerosol transmission of COVID-19 to five fairly well documented cases of viral transmission. The cases were chosen because exposure times, environment volumes, and infection rates are well characterized and publicly available. The simple model assumes that the probability of infection after inhalation of *N* virions is 1 – exp(-*N*/*N*_0_) and that the concentration of viral particles in the room can decay over time due to exchanges of air and to degradation of the virus particles. We also assume that the rate at which the infected person is expelling virions is the product of concentration of viable virions in a volume of exhaled fluid and the volume of fluid exhaled per time.

One of the key differences between our work and prior work is our conclusion that so-called “superspreading” events can be adequately explained by index individuals with normal viral loads, while prior works analyzing some of the cases considered here have estimated the viral loads of index individuals to be higher than typical [20,21,29]. The inference of high viral loads in prior work stems from an assumption of low volumes of aerosols emitted by index patients (order 1-10 nL/hour) [20,21]. In contrast, based on Stadnytskyi’s measurements, we estimate ∼30 μL/hr emitted are emitted when speaking [33]. The literature includes a wide variation in measurements of the volume of exhaled fluid and in the fraction of viable viral particles within that fluid volume [9,10,33]. Further research in those areas would be very helpful in formulating effective guidelines for reducing the spread of COVID-19. However, even absent reliable information on those values our model indicates that talking and breathing release ∼460 *N*_0_ and ∼10*N*_0_ (quanta)/hour, which can allow people to estimate risks of everyday activities even if the values of *N*_0_ are uncertain [20–22,28]. Reducing the uncertainty in the volume of exhaled fluids in the fraction of viable virus particles would be valuable in framing these risks. If, as calculated here, these quanta correspond to fairly typical viral densities, the population-wide risks would be reasonably represented by the range of values calculated here. However, if the five cases here corresponded to index patients with exceptionally high viral loads, the population-wide risks of aerosols could be much lower. Applying published values for the concentration of viable virions in a volume of exhaled fluid and the volume of fluid exhaled per time to our model, we separately obtain *N*_0_ values for each of the five cases. Importantly, we obtain a narrow range of *N*_0_ values (∼300-2000), despite large variations in air volumes and exposure times. It is somewhat remarkable that these consistent results were obtained using a single value of viral load (copies/mL) for an infected person who spreads the virus, even though the measured values for copies/mL vary over 8-9 orders of magnitude. If the range of ϕ(t) is narrow, a narrow range in *N*_0_ is a signature of relatively low dispersion in the viral loads of the index patients considered here. That conclusion is consistent with measurements of viral load as a function of time indicating that peak infectivity occurs during a very short time when the viral load is a maximum. Importantly, the modeled and observed maxima fall within a fairly narrow range [1,2]. Finally, as another check, applying the same values of *N*_0_ to fomite transmission (at an average viral load) suggests fomites might typically provide order 0.01-0.1 *N*_0_ of exposure, consistent with the observation that fomites are likely not a large transmission channel.

In sum, our application of a simple aerosol transmission model to five well characterized transmission events suggests that the viral loads in index patients, and the *N*_0_ values for infection are similar for COVID-19 and influenza, which also has airborne transmission [16,69]; however, COVID-19 transmission is most probable during a very narrow time window (∼1 day) at the onset of symptoms, whereas for many viruses, including influenza, transmission is dominated by symptomatic patients who may remain infectious for days [1,2]. When comparing the spread of COVID-19 with the spread of other viral infections it is important to note that transmission of viruses is greatly reduced if there is significant herd immunity. Thus, we propose that from “enhanced transmission” due to an index patient with an average viral load who is just becoming symptomatic may play a very important role in the COVID-19 pandemic. Transmission by that patient is enhanced because a person or people with no previous exposure to the virus is confined with the index patient in a space that accumulates airborne particles (amplified by poor ventilation, vigorous activity, or lack of masks). Thus, “superspreader” events simply represent extreme examples of enhanced transmission in which many people are simultaneously confined with the index patient. Importantly, we also find that the required confinement period may be as short as an hour and that enhanced transmission can occur at events attended by only a small number of people, such as family dinners. Our findings support many of the existing measures being employed to suppress transmission of COVID-19, including mask wearing; however, the results also suggest that separating people by at least six feet and screening against individuals with significant symptoms may not strongly reduce COVID-19 transmission.

## Data Availability

The data was obtained from publicly available information.

## Acknowledgements and Additional Disclosures

We thank John Doyle, Claudia Danilowicz, James Kwak, James Fujimoto, and Richard Garwin for useful comments / discussions, and Steven Kleiboeker for sharing his viral load data. The labs of Mara Prentiss and Karl Berggren have been recipients of research funding from the Chu Family Foundation, for topics unrelated to this work.

## Footnotes

1 For example, Buannano, Augenbraun, and Miller assume representative values of *N*_0_ = 50, 100, and 1,000 respectively [20,29,30], not dissimilar from the speculations of certain virologists who posit an infective dose of a few hundred to a few thousand particles [31].

2 We note that our analysis of the Skagit choir event finds a similar quanta emission rate to previous work that concluded the event involved a superspreading individual [20]; however, we used a fluid emission rate for the index patient that was much larger than the rate put forth in the previous Skagit choir study (1-10 nL/hour); therefore, since the infectious particle emission rate is the product of the viral load and the fluid emission rate, our much lower estimate of copies/mL was still consistent with the quanta emission rate used in both studies.

3 The source of this discrepancy is unclear, though it is possible that the measurement of van Doremalen includes some effect of settling. Fears *et al* explicitly note that the rotation rate of the Goldberg drum used was sufficiently high to create a suspended aerosol[45]. Unrelated measurements by Huang *et al* found a half life of 1.38 ± 0.05 hours for 3 μm particles rotating at 5 rpm in a Goldberg drum, not dissimilar from the 1.09 hour half life found by van Doremalen[46].

4 The standard deviation for Jones et al is estimated from Table 2 [18]. Kleiboeker *et al* do not report a standard deviation, so the figure of ∼2.0 log_10_ copies/mL was calculated based on the underlying data provided to us by Kleiboeker (private communication). The mean log_10_ copies/mL for Arnaout and Jacot were calculated based on digitizing the data in their figures 2(a) and 1(a), respectively [16,17].

5 Jacot *et al* only measure the initial sample of a given patient, but these patients were tested at different points during disease progression[16].

6 The factor of 17.7 is obtained by comparing the average of the “aah-v-p” and “c-v-p” number concentrations with the average of the “b-n-m” and “b-n-n” concentrations. Note that in the vocalization experiments, equal time is spent speaking and breathing, so we remove the “background” concentration due to the time spent only breathing. The factor of 45.2 is obtained the same way, summing the volume contributions of each measured diameter bucket from 0.8 μm to 5.5 μm.

7 Ma *et al* used EBC to measure SARS-Cov-2 emissions in breath and found positive readings in 14 out of 52 (26%) of the EBC samples. They infer an emission rate of 1.03× 10^5^ - 2.25 × 10^7^ virus copies/hour [48], which is substantially higher than the ∼6,000 copies/hour implied by our breathing calculation. The size of the EBC samples ranges from 300-500 μL for 5 minutes of collection. At a dilution factor of 20,000, the aerosol emission rate would be ∼0.4 mL/5 minutes/20000 ∼ 2.4 × 10^−4^ mL/hour (similar to our value of *ϕ*_breathing_ of 6.0 × 10^−4^ mL/hour), which in turn would imply a viral load of ∼4 × 10^8^ - 9 × 10^10^ copies/mL in the exhaled aerosols. These viral loads are very high (∼95-100% percentile of the viral load ranges measured by Kleiboeker [15]. It is possible that the discrepancy between the values is due to the conversion of cycle threshold (Ct) values to copies/mL; the conversion reported by Ma is 2-4 orders of magnitude higher than that reported by other authors. Ma *et al* convert Ct values to copies/mL according to the formula copies/mL = 10^5^ × 1.75^(39.5-Ct) [48]^, or equivalently, Log_10_ (copies/mL) = 14.60 - 0.24 Ct. For the average Ct value of 35.54 reported by Ma, this conversion implies a viral load in the EBC of ∼6.0 Log_10_ copies/mL. If the respiratory fluids in EBC are diluted by a factor of ∼20,000 by water vapor, the viral load in respiratory fluid would be ∼2 × 10^10^ copies/mL (∼10.3 Log_10_ (copies/mL), at the extreme high of measured values. In contrast, Zou (Log_10_ (copies/mL = 14.11 - 0.32 Ct), Jones (Log_10_ (copies/mL) = 14.16 - 0.30 Ct), Kleiboeker (Log_10_ (copies/mL) = 12.27 - 0.30 Ct, and Jacot (Log_10_ (copies/mL) = 13.14 - 0.27Ct) all would find a much lower viral load in EBC for a Ct value of 35.54 -- 2.6, 3.6, 1.6, and 3.5 Log_10_ (copies/mL), respectively. Increasing these by a factor of 20,000 for EBC dilution would produce a viral load in respiratory fluid of ∼5.9-7.9 Log_10_ copies/mL, which is more in line with measured distributions [15,16,18,49].

8 Although the individual reduction factor measurements vary widely, the dilution estimates are much tighter, at 20,472 ± 2,516 for cation measurements, 21,019 ± 2,427 for conductivity measurements, and 18,818 ± 2,402 for urea measurements.

9 In a later paper, Effros et al cite a dilution factor of approximately 10,000 [35].

10 In the case of the Skagit choir, based on a more detailed calculation, Miller *et al* use a smaller air exchange of 0.3-1.0 h^-1^. However, adding the contribution of ventilation loss rates deposition, and virus inactivation, the average total decay rate in their calculation is 1.9 h^-1^, which is similar to the 2.1 h^-1^ used here [20].

11 The event in the bus occurred during winter, so the bus windows were believed to be closed (Y. Shen, personal communication).

12 In the case of the Wuhan buses, the index patient is thought to have had a mild cough (Y. Yang, personal communication).

13 Note that in the Skagit choir case, the calculated quanta emission is of 2,347/h is roughly double the mean emission rate of 970 quanta/h reported by Miller *et al* [20]; this difference is largely due to a mean assumed breathing rate in the Miller calculation of ∼1.0 m^3^/h (range 0.65-1.38), compared to 0.5 m^3^/h here.

14 For the Korean call center case, there is uncertainty about the time that the index patient spent in the call center. We have assumed 8 hours (1 full day), but it is possible that the index patient was present for more than 1 day. We note that using 16 hours, gives *N*_0_ = 745 and using 24 hours gives *N*_0_ = 1,129, so our results are robust over a range of reasonable exposure times.

15 Using Figure 2 of Fears *et al*, the genome/PFU ratio is estimated to be ∼20, ∼1040, ∼1140, ∼189, and ∼132 after 10, 30, 120, 240, and 960 minutes, respectively.[45] Buonanno *et al* appear use the “steady state” figure of 1.3 × 10^2^ for c_PFU_ (copies per PFU) in their analysis [22].

16 The value of ∼500 genome copies/PFU is in the range of that measured for SARS-CoV-1, where Vicenzi *et al* measured 360 genome copies/PFU [63]. However, Watanabe *et al* estimated that the ID50 for SARS-CoV-1 is estimated to be 280 PFU (*N*_*0*_ = 410 PFU), implying that SARS-CoV-1 would have *N*_*0*_ of order 150,000 genome copies.

17 It should be noted that this range is broadened by measurement differences: the data contains 9 patients from Germany which were analyzed using sputum samples. These samples are systematically higher than those from other 16 patients, which were analyzed using nasopharyngeal (NP) swabs. The modeled peak values for the sputum samples were in the range of ∼8.2-8.3 log_10_ copies/sample, while the modeled peak values for the NP samples were in the range of ∼7.2-7.3 log_10_ copies/sample.

18 As in the case of the modeled distributions, the sputum samples generally show higher viral loads than the NP samples. The data at 8.5 and 9.0 log_10_ copies are from sputum samples.

19 Jacot notes that the distributions for these other viruses are statistically significantly different in 9 of the 14 cases (Jacot Figure 2); we are simply noting that the medians across viruses, even if different statistically, are of similar magnitudes.

20 There is a question of how to model the effect of filtration and HVAC systems generally. While one might worry that an HVAC system would spread the virus across floors of a building, there have been no documented cases of this occurring. To the contrary, as a virion makes its way through a complicated maze of ductwork, it encounters various surfaces, each of which has some probability of capturing the virus or otherwise inactivating it/rendering it non-infectious. Thus, one can make a cogent argument that an HVAC system could create some attenuation factor, possibly much smaller than 1, which would lead to a more powerful viral attenuation than just rated value of its filter (i.e. more powerful than the -*R* ln(*Q*) contribution above). This question of coronavirus attenuation due to mechanical airflow is a subject that, to the best of our knowledge, has not been studied quantitatively. To be conservative, we assume there is no attenuation beyond what is in the rated value of the filter.

21 The well-mixed approximation is likely not strictly correct in all cases. For example, the volume of the supermarket is so large that the timescales do not strictly justify the well-mixed approximation over the entire volume; however, even if one used a much smaller volume, the risks would still be very low. Similarly, in the case of an airplane, there is active airflow and recirculation which may create somewhat distinct, smaller zones similar to those observed in the Guangzhou restaurant case.

22 Given this small exposure rate and the ubiquity of people touching their eyes, nose, or mouth, one immediately wonders if fomite transmission of sub-single-quantum doses of virions through touching of contaminated surfaces and subsequent inadvertent inoculation of the mucosal membranes by face touching might play a role in protection from infectious disease through natural variolation. A similar effect has been suggested to occur if a low volume of virions are transmitted between masked individuals when social distancing is being practiced.[68]

## REFERENCES

1. Goyal A, Cardozo-Ojeda EF, Schiffer JT. Potency and timing of antiviral therapy as determinants of duration of SARS CoV-2 shedding and intensity of inflammatory response. Infectious Diseases (except HIV/AIDS). medRxiv; 2020. doi:10.1101/2020.04.10.20061325

2. Goyal A, Reeves DB, Cardozo-Ojeda EF, Schiffer JT, Mayer BT. Wrong person, place and time: viral load and contact network structure predict SARS-CoV-2 transmission and super-spreading events. Infectious Diseases (except HIV/AIDS). medRxiv; 2020. doi:10.1101/2020.08.07.20169920

3. Lewis D. Mounting evidence suggests coronavirus is airborne - but health advice has not caught up. Nature. 2020. pp. 510–513.

4. Tellier R, Li Y, Cowling BJ, Tang JW. Recognition of aerosol transmission of infectious agents: a commentary. BMC Infect Dis. 2019;19: 101.

5. Cascella M, Rajnik M, Cuomo A, Dulebohn SC, Di Napoli R. Features, Evaluation, and Treatment of Coronavirus (COVID-19). StatPearls Publishing; 2020.

6. Paul Baron, Division of Applied Technology, NIST and CDC. Generation and Behavior of Airborne Particles. Available: https://www.cdc.gov/niosh/topics/aerosols/pdfs/Aerosol_101.pdf

7. Papineni RS, Rosenthal FS. The size distribution of droplets in the exhaled breath of healthy human subjects. J Aerosol Med. 1997;10: 105–116.

8. Zayas G, Chiang MC, Wong E, MacDonald F, Lange CF, Senthilselvan A, et al. Cough aerosol in healthy participants: fundamental knowledge to optimize droplet-spread infectious respiratory disease management. BMC Pulm Med. 2012;12: 11.

9. Asadi S, Wexler AS, Cappa CD, Barreda S, Bouvier NM, Ristenpart WD. Aerosol emission and superemission during human speech increase with voice loudness. Sci Rep. 2019;9: 2348.

10. Morawska L, Johnson GR, Ristovski ZD, Hargreaves M, Mengersen K, Corbett S, et al. Size distribution and sites of origin of droplets expelled from the human respiratory tract during expiratory activities. J Aerosol Sci. 2009;40: 256–269.

11. Lednicky JA, Lauzardo M, Hugh Fan Z, Jutla AS, Tilly TB, Gangwar M, et al. Viable SARS-CoV-2 in the air of a hospital room with COVID-19 patients. medRxiv. 2020;2020.08.03.20167395.

12. How COVID-19 Spreads. In: CDC website [Internet]. Oct 2020 [cited Oct 2020]. Available: https://www.cdc.gov/coronavirus/2019-ncov/prevent-getting-sick/how-covid-spreads.html#edn1

13. Adam DC, Wu P, Wong JY, Lau EHY, Tsang TK, Cauchemez S, et al. Clustering and superspreading potential of SARS-CoV-2 infections in Hong Kong. Nat Med. 2020. doi:10.1038/s41591-020-1092-0

14. Lau MSY, Grenfell B, Thomas M, Bryan M, Nelson K, Lopman B. Characterizing superspreading events and age-specific infectiousness of SARS-CoV-2 transmission in Georgia, USA. Proc Natl Acad Sci U S A. 2020;117: 22430–22435.

15. Kleiboeker S, Cowden S, Grantham J, Nutt J, Tyler A, Berg A, et al. SARS-CoV-2 viral load assessment in respiratory samples. J Clin Virol. 2020;129: 104439.

16. Jacot D, Greub G, Jaton K, Opota O. Viral load of SARS-CoV-2 across patients and compared to other respiratory viruses. Infectious Diseases (except HIV/AIDS). medRxiv; 2020. doi:10.1101/2020.07.15.20154518

17. Arnaout R, Lee RA, Lee GR, Callahan C, Yen CF, Smith KP, et al. SARS-CoV2 Testing: The Limit of Detection Matters. bioRxiv. 2020. doi:10.1101/2020.06.02.131144

18. Jones TC, Mühlemann B, Veith T, Biele G, Zuchowski M, Hoffmann J, et al. An analysis of SARS-CoV-2 viral load by patient age. Infectious Diseases (except HIV/AIDS). medRxiv; 2020. doi:10.1101/2020.06.08.20125484

19. Wölfel R, Corman VM, Guggemos W, Seilmaier M, Zange S, Müller MA, et al. Virological assessment of hospitalized patients with COVID-2019. Nature. 2020;581: 465–469.

20. Miller SL, Nazaroff WW, Jimenez JL, Boerstra A, Buonanno G, Dancer SJ, et al. Transmission of SARS-CoV-2 by inhalation of respiratory aerosol in the Skagit Valley Chorale superspreading event. Infectious Diseases (except HIV/AIDS). medRxiv; 2020. doi:10.1101/2020.06.15.20132027

21. Bazant MZ, Bush JWM. Beyond Six Feet: A Guideline to Limit Indoor Airborne Transmission of COVID-19. medRxiv. 2020. Available: https://www.medrxiv.org/content/10.1101/2020.08.26.20182824v1.abstract

22. Buonanno G, Morawska L, Stabile L. Quantitative assessment of the risk of airborne transmission of SARS-CoV-2 infection: prospective and retrospective applications. Environment International. 2020. p. 106112. doi:10.1016/j.envint.2020.106112

23. Shen Y, Li C, Dong H, Wang Z, Martinez L, Sun Z, et al. Community Outbreak Investigation of SARS-CoV-2 Transmission Among Bus Riders in Eastern China. JAMA Intern Med. 2020. doi:10.1001/jamainternmed.2020.5225

24. Luo K, Lei Z, Hai Z, Xiao S, Rui J, Yang H, et al. Transmission of SARS-CoV-2 in Public Transportation Vehicles: A Case Study in Hunan Province, China. Open Forum Infect Dis. 2020 [cited 25 Sep 2020]. doi:10.1093/ofid/ofaa430

25. Jang S, Han SH, Rhee J-Y. Cluster of Coronavirus Disease Associated with Fitness Dance Classes, South Korea. Emerg Infect Dis. 2020;26: 1917–1920.

26. Park SY, Kim Y-M, Yi S, Lee S, Na B-J, Kim CB, et al. Coronavirus Disease Outbreak in Call Center, South Korea. Emerg Infect Dis. 2020;26: 1666–1670.

27. Hamner L, Dubbel P, Capron I, Ross A, Jordan A, Lee J, et al. High SARS-CoV-2 Attack Rate Following Exposure at a Choir Practice - Skagit County, Washington, March 2020. MMWR Morb Mortal Wkly Rep. 2020;69: 606–610.

28. Jimenez JL. COVID-19 Aerosol Transmission Estimator. [cited Sep 2020]. Available: https://tinyurl.com/covid-estimator

29. Buonanno G, Stabile L, Morawska L. Estimation of airborne viral emission: Quanta emission rate of SARS-CoV-2 for infection risk assessment. Environ Int. 2020;141: 105794.

30. B. L. Augenbraun, Z. D. Lasner, D. Mitra, S. Prabhu, S. Raval, H. Sawaoka, J. M. Doyle. Assessment and Mitigation of Aerosol Airborne SARS-CoV-2 Transmission in Laboratory and Office Environments. J Occup Environ Hyg. 2020.

31. “Expert Reactions to Questions About COVID-19 and viral load.” Mar 2020 [cited Sep 2020]. Available: https://www.sciencemediacentre.org/expert-reaction-to-questions-about-covid-19-and-viral-load/

32. Yang W, Elankumaran S, Marr LC. Concentrations and size distributions of airborne influenza A viruses measured indoors at a health centre, a day-care centre and on aeroplanes. J R Soc Interface. 2011;8: 1176–1184.

33. Stadnytskyi V, Bax CE, Bax A, Anfinrud P. The airborne lifetime of small speech droplets and their potential importance in SARS-CoV-2 transmission. Proc Natl Acad Sci U S A. 2020;117: 11875–11877.

34. Effros RM, Biller J, Foss B, Hoagland K, Dunning MB, Castillo D, et al. A simple method for estimating respiratory solute dilution in exhaled breath condensates. Am J Respir Crit Care Med. 2003;168: 1500–1505.

35. Effros RM, Casaburi R, Porszasz J, Morales EM, Rehan V. Exhaled breath condensates: analyzing the expiratory plume. Am J Respir Crit Care Med. 2012;185: 803–804.

36. Effros RM, Dunning MB 3rd, Biller J, Shaker R. The promise and perils of exhaled breath condensates. Am J Physiol Lung Cell Mol Physiol. 2004;287: L1073–80.

37. Ferretti L, Wymant C, Kendall M, Zhao L, Nurtay A, Abeler-Dörner L, et al. Quantifying SARS-CoV-2 transmission suggests epidemic control with digital contact tracing. Science. 2020;368. doi:10.1126/science.abb6936

38. Sze To GN, Chao CYH. Review and comparison between the Wells-Riley and dose-response approaches to risk assessment of infectious respiratory diseases. Indoor Air. 2010;20: 2–16.

39. Shimer DA, Jenkins PL, Hui SP, Adams WC. 132 MEASUREMENT OF BREATHING RATE AND VOLUME IN ROUTINELY PERFORMED DAILY ACTIVITIES. Epidemiology. 1995;6: S30.

40. Binazzi B, Lanini B, Bianchi R, Romagnoli I, Nerini M, Gigliotti F, et al. Breathing pattern and kinematics in normal subjects during speech, singing and loud whispering. Acta Physiol. 2006;186: 233–246.

41. U.S. EPA Office of Research and Development. Chapter 6 (Inhalation Rates). Exposure Factors Handbook.

42. Lindsley WG, King WP, Thewlis RE, Reynolds JS, Panday K, Cao G, et al. Dispersion and exposure to a cough-generated aerosol in a simulated medical examination room. J Occup Environ Hyg. 2012;9: 681–690.

43. Diapouli E, Chaloulakou A, Koutrakis P. Estimating the concentration of indoor particles of outdoor origin: a review. J Air Waste Manag Assoc. 2013;63: 1113–1129.

44. van Doremalen N, Bushmaker T, Morris DH, Holbrook MG, Gamble A, Williamson BN, et al. Aerosol and Surface Stability of SARS-CoV-2 as Compared with SARS-CoV-1. N Engl J Med. 2020;382: 1564–1567.

45. Fears AC, Klimstra WB, Duprex P, Hartman A, Weaver SC, Plante KC, et al. Comparative dynamic aerosol efficiencies of three emergent coronaviruses and the unusual persistence of SARS-CoV-2 in aerosol suspensions. medRxiv. 2020. doi:10.1101/2020.04.13.20063784

46. Huang S-H, Kuo Y-M, Lin C-W, Ke W-R, Chen C-C. Experimental Characterization of Aerosol Suspension in a Rotating Drum. Aerosol Air Qual Res. 2019;19: 688–697.

47. Loudon RG, Roberts RM. Droplet expulsion from the respiratory tract. Am Rev Respir Dis. 1967;95: 435–442.

48. Ma J, Qi X, Chen H, Li X, Zhang Z, Wang H, et al. COVID-19 patients in earlier stages exhaled millions of SARS-CoV-2 per hour. Clin Infect Dis. 2020. doi:10.1093/cid/ciaa1283

49. Zou L, Ruan F, Huang M, Liang L, Huang H, Hong Z, et al. SARS-CoV-2 Viral Load in Upper Respiratory Specimens of Infected Patients. N Engl J Med. 2020;382: 1177–1179.

50. Brandis K. Insensible Water Loss, Section 3.2 of Fluid Physiology. [cited Oct 2020]. Available: https://paperpile.com/app

51. Thomas RJ. Particle size and pathogenicity in the respiratory tract. Virulence. 2013;4: 847–858.

52. Santarpia JL, Herrera VL, Rivera DN, Ratnesar-Shumate S, Denton PW, Martens JWS, et al. The infectious nature of patient-generated sars-cov-2 aerosol. medRxiv. 2020. Available: https://www.medrxiv.org/content/10.1101/2020.07.13.20041632v2.abstract

53. Thatcher TL, Lai ACK, Moreno-Jackson R, Sextro RG, Nazaroff WW. Effects of room furnishings and air speed on particle deposition rates indoors. Atmos Environ. 2002;36: 1811–1819.

54. Cox J. CHORAL REHEARSAL TIME USAGE IN A HIGH SCHOOL AND A UNIVERSITY: A COMPARATIVE ANALYSIS. Contributions to Music Education. 1986; 7–22.

55. Liu Y, Ning Z, Chen Y, Guo M, Liu Y, Gali NK, et al. Aerodynamic analysis of SARS-CoV-2 in two Wuhan hospitals. Nature. 2020;582: 557–560.

56. Santarpia JL, Rivera DN, Herrera V, Morwitzer MJ, Creager H, Santarpia GW, et al. Aerosol and Surface Transmission Potential of SARS-CoV-2. Infectious Diseases (except HIV/AIDS). medRxiv; 2020. doi:10.1101/2020.03.23.20039446

57. Killingley B, Greatorex J, Digard P, Wise H, Garcia F, Varsani H, et al. The environmental deposition of influenza virus from patients infected with influenza A(H1N1)pdm09: Implications for infection prevention and control. J Infect Public Health. 2016;9: 278–288.

58. Bischoff WE, Swett K, Leng I, Peters TR. Exposure to influenza virus aerosols during routine patient care. J Infect Dis. 2013;207: 1037–1046.

59. Alford RH, Kasel JA, Gerone PJ, Knight V. Human influenza resulting from aerosol inhalation. Proc Soc Exp Biol Med. 1966;122: 800–804.

60. Fabian P, McDevitt JJ, DeHaan WH, Fung ROP, Cowling BJ, Chan KH, et al. Influenza virus in human exhaled breath: an observational study. PLoS One. 2008;3: e2691.

61. Ward C. Design and performance testing of quantitative real time PCR assays for influenza A and B viral load measurement. Journal of Clinical Virology. 2004. pp. 179–188. doi:10.1016/s1386-6532(03)00122-7

62. Converting TCID[50] to plaque forming units (PFU). In: ATCC FAQ [Internet]. [cited Sep 2020]. Available: https://www.atcc.org/support/faqs/48802/Converting%20TCID50%20to%20plaque%20forming%20units%20PFU-124.aspx

63. Vicenzi E, Canducci F, Pinna D, Mancini N, Carletti S, Lazzarin A, et al. Coronaviridae and SARS-associated coronavirus strain HSR1. Emerg Infect Dis. 2004;10: 413–418.

64. He X, Lau EHY, Wu P, Deng X, Wang J, Hao X, et al. Temporal dynamics in viral shedding and transmissibility of COVID-19. Nat Med. 2020;26: 672–675.

65. Milton DK, Fabian MP, Cowling BJ, Grantham ML, McDevitt JJ. Influenza virus aerosols in human exhaled breath: particle size, culturability, and effect of surgical masks. PLoS Pathog. 2013;9: e1003205.

66. Gou Y. COVID 19 Projections Website. Oct 2020 [cited Oct 2020]. Available: https://covid19-projections.com/us

67. Zhang N, Li Y. Transmission of Influenza A in a Student Office Based on Realistic Person-to-Person Contact and Surface Touch Behaviour. Int J Environ Res Public Health. 2018;15. doi:10.3390/ijerph15081699

68. Gandhi M, Rutherford GW. Facial Masking for Covid-19 - Potential for “Variolation” as We Await a Vaccine. N Engl J Med. 2020. doi:10.1056/NEJMp2026913

69. Tellier R. Review of aerosol transmission of influenza A virus. Emerg Infect Dis. 2006;12: 1657–1662.

