## Supplemental Information for "Superspreading Events Without Superspreaders: Using High Attack Rate Events to Estimate N_º_ for Airborne Transmission of COVID-19"

### S1 Representative Values of Air Exchange Rate $\lambda_1$

The air exchange rate $\lambda_1$ varies significantly depending on the setting.

**Residential homes** tend to be quite "tight", meaning that air turns over only every few or several hours: the EPA recommends using $\lambda_1$ = 0.45/h (i.e. the air turns over every ~1/0.45 ~ 2 hours) for residential homes broadly, while the California Department of Public Health recommends using $\lambda_1$ = 0.23/h based on measurements in newer homes, which tend to have better insulation and less air exchange with the outside [1].

**Office buildings and institutional buildings** usually have higher air exchange rates – circa 1.0/h in schools, dormitories, etc[2]; the EPA recommends a mean value of 1.5/h for nonresidential buildings [3].

**Motor vehicles** tend to have high air exchange rates. A relatively modern car with the windows closed and vent off was measured to have an ACH of ~1.8/h when stationary, 5.6/h at 35 mph, and 13.5 at 55 mph. With higher outside ventilation, these figures climb to 10.7/h, 35.7/h, and 55/h [4,5]. Measurements when vehicles are traveling with air in "recirculate" mode (as in the winter) find that when vehicles are traveling 60 km/h (~37 mph), ACH range from ~2.5 to ~5.5.[6] Note, however, that in tour buses, particularly those with air set to recirculation mode, the air exchange rates have been characterized as "severely lacking," so that buses may have significantly lower ACH if the windows are closed and the vents are closed [7].

**Airplanes** have 20-30 air exchanges per hour, about half of which is recirculated air, and the half of which is outside air. Note that the recirculated air generally will pass through a strong filter (e.g. a HEPA filter, which eliminates ~99+% of viruses per pass (i.e. is very similar to fresh air) [8]. American Airlines, for instance, claims that the cabin air recirculates every 2-4 minutes through a HEPA filter which eliminates 99.97% of particles, including viral particles [9].

### S2 Case of Guangzhou Restaurant

In the spreadsheet, we also consider a well-known case of a restaurant in Guangzhou, where active air conditioning appeared to spread the virus from an index patient to nearby tables in the recirculation zone [10]. One strength of this case is that the airflow characteristics are well-known and well-modeled. Li *et al* have analyzed this case in detail as a probable case of aerosol transmission, including performing tracer gas measurements to find an average air exchange rate of ~0.67 $h^{-1}$ and performing detailed computational fluid dynamics simulations [11]. However, the attack rate is not well defined: 3 out of 4 people in one family, and 2 out of 7 people in another family (both families in the recirculation zone) were infected, but it is unknown how many of these infections were due to exposure at the restaurant.

Lu *et al* note that it is likely that all people were infected at the restaurant but that it is also possible that family transmission played a role, so the attack rate ranges from 2/11 to 5/11 [10]. In addition, it is unclear what volume to apply in a simple well-mixed model. The air conditioned zone is not well-mixed with the other spaces in the restaurant but nor is it clearly a separate zone with aerosol concentrations that dominate the other spaces: Li *et al*. calculate and measure (in their table S1) that the air conditioned zone where the infections took place have an tracer gas density 2-3 times that in more remote regions of the restaurant, where no patrons were infected [11]. Taking all of these uncertainties into account, we can still perform a stylized calculation assuming that the relevant volume is either the zone of the air conditioning or the entire restaurant. We find a similar range of $N_0$ = 499-948 if we assume all infected individuals were infected in the restaurant, and a range of $N_0$ = 1,507-2,415 if only one member of each family was infected at the restaurant.

REFERENCES


1. Chen W, Persily AK, Hodgson AT, Offermann FJ, Poppendieck D, Kumagai K. Area-specific airflow rates for evaluating the impacts of VOC emissions in U.S. single-family homes. Build Environ. 2014;71: 204–211.

2. You Y, Niu C, Zhou J, Liu Y, Bai Z, Zhang J, et al. Measurement of air exchange rates in different indoor environments using continuous CO2 sensors. J Environ Sci . 2012;24: 657–664.

3. U.S. EPA Office of Research and Development. Chapter 19 (Building Characteristics). Exposure Factors Handbook.

4. Ott W, Klepeis N, Switzer P. Air change rates of motor vehicles and in-vehicle pollutant concentrations from secondhand smoke. J Expo Sci Environ Epidemiol. 2008;18: 312–325.

5. Rodes C, Sheldon L, Whitaker D, Clayton A, Fitzgerald K, Flanagan J, et al. Measuring concentrations of selected air pollutants inside California vehicles. 1998. Available: https://rosap.ntl.bts.gov/view/dot/15680/

6. Tong Z, Li Y, Westerdahl D, Adamkiewicz G, Spengler JD. Exploring the effects of ventilation practices in mitigating in-vehicle exposure to traffic-related air pollutants in China. Environ Int. 2019;127: 773–784.

7. Chiu C-F, Chen M-H, Chang F-H. Carbon Dioxide Concentrations and Temperatures within Tour Buses under Real-Time Traffic Conditions. PLoS One. 2015;10: e0125117.

8. Elwood H. Hunt DRS. The Airplane Cabin Environment: Issues Pertaining to Flight Attendant. 1994. Available: http://citeseerx.ist.psu.edu/viewdoc/summary?doi=10.1.1.304.7321

9. American Airlines. How HEPA Filters Have Been Purifying Cabin Air Since the 1990s. In: American Airlines website, News Releases [Internet]. 26 Jun 2020 [cited Sep 2020]. Available: https://news.aa.com/news/news-details/2020/How-HEPA-Filters-Have-Been-Purifying-Cabin-Air-Since-the-1990s-FLT-06/default.aspx#:~:text=The%20HEPA%20filters%20in%20use,engine%20compressor%20on%20the%20outside.

10. Lu J, Gu J, Li K, Xu C, Su W, Lai Z, et al. COVID-19 Outbreak Associated with Air Conditioning in Restaurant, Guangzhou, China, 2020. Emerging Infectious Diseases. 2020. pp. 1628–1631. doi:10.3201/eid2607.200764

11. Li Y, Qian H, Hang J, Chen X, Hong L, Liang P, et al. Evidence for probable aerosol transmission of SARS-CoV-2 in a poorly ventilated restaurant. doi:10.1101/2020.04.16.20067728
